# A Deterministic–Stochastic Model for COVID-19 and Malaria Co-Infection with Malaria-Acquired Partial Immunity

**DOI:** 10.64898/2026.04.27.26351858

**Authors:** Kabir Oluwatobi Idowu, Guang Lin

**Affiliations:** Department of Mathematics, Purdue University, West Lafayette 47907, Indiana, USA; School of Mechanical Engineering, Purdue University, West Lafayette 47907, Indiana, USA

## Abstract

Coinfection of COVID-19 and malaria in endemic regions may generate complex epidemiological interactions that influence susceptibility patterns, disease burden, and outbreak risk. Although malaria-acquired immunity has been hypothesized to modulate host responses to other infections, its population-level implications for COVID-19 transmission under uncertainty remain insufficiently understood. In this study, we develop a deterministic-stochastic compartmental model for the coupled dynamics of COVID-19, malaria, and their co-infection. Malaria-acquired partial immunity is incorporated through a relative susceptibility parameter that reduces the risk of COVID-19 infection among malaria-recovered individuals.

For the deterministic system, we establish positivity, boundedness, an invariant feasible region, and basic reproduction numbers for the COVID-19-only and malaria-only subsystems. We then use numerical simulations to examine how immunity-mediated reductions in susceptibility may influence COVID-19 incidence, peak burden, hospitalization, and cumulative mortality. To account for environmental and transmission variability, we extend the deterministic model to an Itô stochastic differential equation framework and use repeated realizations to characterize uncertainty in epidemic trajectories, peak distributions, and outbreak risk. In addition, global sensitivity analysis based on partial rank correlation coefficients (PRCCs) is performed to identify the parameters with the greatest influence on COVID-19 outcomes.

Our results suggest that, under the assumed modeling framework, malaria-acquired partial immunity may reduce the peak infectious burden and cumulative mortality associated with COVID-19. The stochastic simulations further show substantial variability around deterministic trajectories and indicate a non-negligible probability of large outbreak events that are not fully captured by mean-field predictions alone. Overall, the proposed framework provides an uncertainty-aware, mechanistic basis for studying COVID-19–malaria co-dynamics and for assessing how interacting disease processes may shape epidemic outcomes in endemic settings.

## Introduction

Malaria and Coronavirus Disease 2019 (COVID-19) remain important public health concerns, particularly in low- and middle-income countries where health systems are often fragile [1–3]. Malaria, a mosquito-borne parasitic disease caused by *Plasmodium* species, continues to impose a substantial burden, especially in sub-Saharan Africa [4]. Despite decades of control efforts, malaria still contributes considerably to morbidity and mortality because of persistent transmission, limited access to care, and the continued emergence of antimalarial drug resistance [5]. In parallel, the COVID-19 pandemic, caused by severe acute respiratory syndrome coronavirus 2 (SARS-CoV-2), placed major strain on health systems worldwide, disrupted routine control programs, and intensified the burden of endemic infectious diseases [6, 7].

In malaria-endemic regions, the interaction between COVID-19 and malaria has received growing attention [8–11]. Clinically, the two diseases share several symptoms, including fever, headache, fatigue, myalgia, and, in some cases, respiratory distress (Fig. 1), which complicates differential diagnosis and may delay appropriate treatment [11–13]. In settings with limited diagnostic capacity, this overlap may increase misclassification, underreporting, and missed co-infections. In addition, documented cases of concurrent malaria and COVID-19 infection motivate a closer examination of how the two pathogens may interact at both the individual and population levels [11].

**Fig 1.**
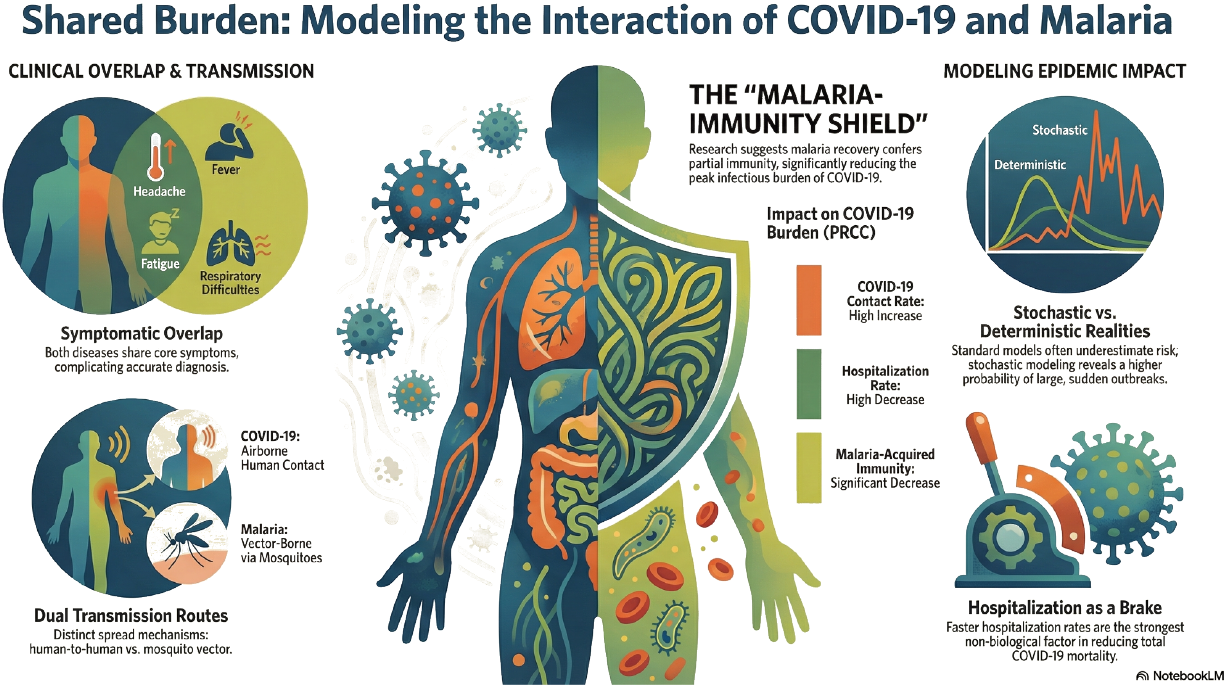
COVID-19 and malaria share several clinical symptoms, which may complicate differential diagnosis in endemic settings.

Beyond clinical overlap, a growing body of literature suggests that malaria exposure may influence COVID-19 susceptibility, severity, or mortality through immunological and genetic mechanisms. Previous studies have identified similarities between malaria and COVID-19 at both innate and adaptive immune levels, including activation of pattern-recognition pathways, cytokine-mediated inflammatory responses, and antibody-related immune processes [14–17]. In particular, Konozy et al. [11] proposed that repeated malaria exposure may contribute to trained immunity and immune tolerance, which could partly influence host responses to SARS-CoV-2 infection in endemic regions. Likewise, Iesa et al. [18] reported potential shared immunodominant regions between SARS-CoV-2 and *Plasmodium falciparum*, suggesting a possible basis for immune cross-reactivity. These findings are suggestive, but they do not yet provide a direct population-level quantification of how malaria-acquired immune history may alter COVID-19 transmission dynamics.

Mathematical modeling provides a useful framework for translating such biological hypotheses into population-level epidemiological questions. Several models have been proposed to study COVID-19–malaria co-infection dynamics. Tchoumi et al. [19] formulated a compartmental model that includes malaria-only, COVID-19-only, and co-infection states in human and mosquito populations, and used optimal control analysis to assess intervention strategies. Although that framework offers useful insight into transmission interactions and control, it does not account for malaria-acquired immunity or stochastic variability in transmission. Abioye et al. [20] extended this line of work by introducing recovered classes and fractional-order derivatives to represent memory effects. However, the model remains deterministic and does not address uncertainty or immunity-mediated changes in COVID-19 susceptibility.

Similarly, Shah et al. [21] studied malaria–COVID-19 co-infection using a deterministic model with distinct incidence functions for the two diseases and focused primarily on control strategies. Avusuglo et al. [22] examined the bidirectional effects of stigmatization-induced self-medication in a deterministic co-infection model for Nigeria. While these studies provide important insight into transmission and control, they do not explicitly incorporate malaria-acquired partial immunity, nor do they quantify the effects of stochastic perturbations arising from environmental, behavioral, or demographic variability. More broadly, existing models have largely emphasized deterministic dynamics and control optimization while giving less attention to uncertainty, immune modulation from repeated malaria exposure, and the role of resistance-driven malaria persistence in shaping co-infection outcomes. Consequently, the population-level implications of malaria-acquired partial immunity for COVID-19 burden and outbreak risk remain insufficiently explored.

Motivated by these gaps, we develop a deterministic–stochastic compartmental model for the coupled transmission of COVID-19, malaria, and their co-infection. The framework explicitly incorporates human–mosquito transmission, hospitalization, disease-induced mortality, drug-sensitive and drug-resistant malaria infection, and a malaria-acquired partial immunity mechanism that reduces susceptibility to COVID-19 among malaria-recovered individuals. To capture both average epidemic behavior and random perturbations in transmission, the deterministic system is extended to an Itô stochastic differential equation model.

For the deterministic formulation, we establish positivity, boundedness, and an invariant feasible region, and we derive the basic reproduction numbers for the COVID-19-only and malaria-only subsystems. We then use numerical simulations to investigate how immunity-mediated reductions in susceptibility may affect COVID-19 incidence, peak burden, hospitalization, and cumulative mortality. To complement these analyses, we incorporate stochastic simulations to characterize variability in epidemic trajectories and outbreak risk, and we perform global sensitivity analysis to identify the parameters that most strongly influence COVID-19 outcomes. In this way, the study provides a mechanistic and uncertainty-aware framework for examining COVID-19–malaria co-dynamics in endemic settings.

## Model Formulation and Diagram

### Model Formulation

The human population is stratified into epidemiological compartments representing the transmission and progression dynamics of COVID-19, malaria, and their co-infection. Susceptible individuals (*S*) are at risk of contracting either infection. They acquire COVID-19 after effective contact with infectious individuals (*I*_*c*_, *I*_*mc*_), moving into the exposed class (*E*_*c*_) at rate *λ*_*c*_. Alternatively, they acquire malaria following the bite of an infectious mosquito, entering either the drug-sensitive malaria class (*I*_*ms*_) with probability (1 − *r*) or the drug-resistant malaria class (*I*_*mr*_) with probability *r*, at rate *λ*_*m*_. Individuals in the COVID-19 exposed class (*E*_*c*_) progress to the infectious stage (*I*_*c*_) at rate *β*_*c*_. Infectious COVID-19 individuals may be hospitalized at a rate *α*_1_ or die directly at a rate *α*_2_. Hospitalized individuals (*H*_*c*_) recover at rate *γ*_1_ or die at rate *d*_1_, with recovery leading to the COVID-recovered class (*R*_*c*_) and death entering the cumulative COVID death class (*D*_*c*_). For malaria dynamics, individuals infected with the drug-sensitive strain (*I*_*ms*_) move to the treated sensitive class (*H*_*ms*_) at rate *b*_1_, while those infected with the drug-resistant strain (*I*_*mr*_) move to the treated resistant class (*H*_*mr*_) at rate *b*_2_ or die directly at rate *b*_3_. Treated individuals recover at rates *ψ*_1_ and *ψ*_2_ for the sensitive and resistant strains, respectively. Severe resistant malaria cases may additionally die at a rate *d*. All malaria recoveries flow into the recovered malaria class (*R*_*m*_), from which partial immunity wanes at rate *ω*_*m*_, returning individuals to the susceptible class (*S*). The model diagram is presented in Fig. 2.

**Fig 2.**
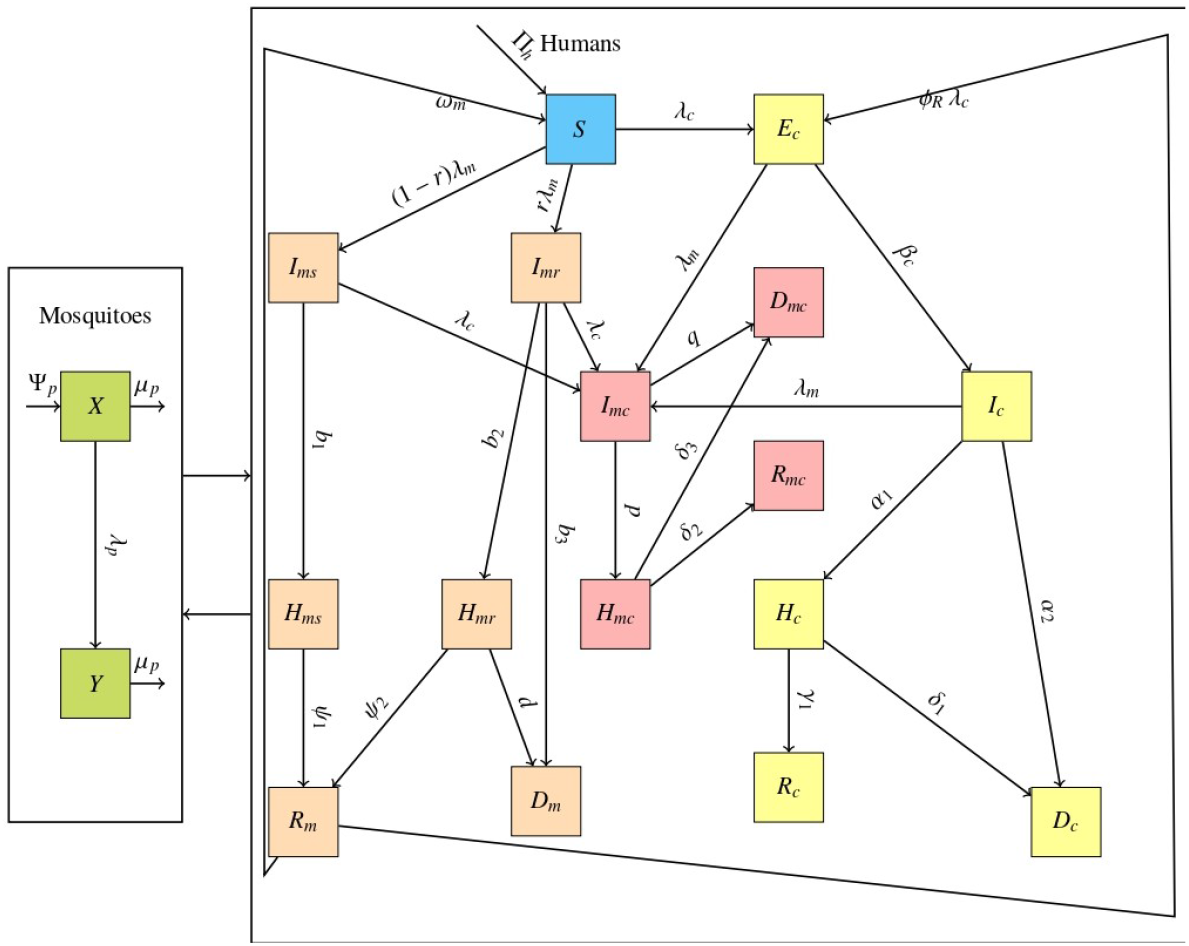
COVID-malaria co-infection model with key transition parameters.

A key interaction mechanism arises through bidirectional acquisition of the two infections. Individuals already infected with malaria (*I*_*ms*_, *I*_*mr*_) may acquire COVID-19 at rate *λ*_*c*_ and transition to the co-infected class (*I*_*mc*_). Conversely, individuals infected with COVID-19 (*E*_*c*_, *I*_*c*_) may acquire malaria at rate *λ*_*m*_ and also transition to (*I*_*mc*_). Thus, co-infection results from the sequential acquisition of the second pathogen by individuals currently infected with the first. Co-infected individuals (*I*_*mc*_) may be hospitalized at rate *p*, entering the hospitalized co-infection class (*H*_*mc*_), or may die directly at rate *q*. Hospitalized co-infected individuals recover at rate *δ*_2_ or die at rate *d*_3_, with survivors entering the recovered co-infection class (*R*_*mc*_). Individuals in (*R*_*mc*_) retain partial malaria-related immune modulation and contribute to mosquito infection at a reduced rate. For parsimony, waning of malaria-acquired immunity is applied only to individuals recovered from malaria-only infection (*R*_*m*_). Over the simulation horizon considered, the post–co-infection recovered class (*R*_*mc*_) is assumed to retain malaria-related immune effects, and no additional waning term is included for *R*_*mc*_. Malaria-acquired partial protection against COVID-19 is incorporated explicitly.

Individuals in the malaria-recovered class (*R*_*m*_) may acquire COVID-19 at a reduced rate *ϕ*_*R*_*λ*_*c*_, where *ϕ*_*R*_ ∈ [0, 1] represents relative susceptibility. They therefore move into the exposed COVID class (*E*_*c*_) at rate *ϕ*_*R*_*λ*_*c*_*R*_*m*_. When *ϕ*_*R*_ = 0, malaria recovery confers complete protection against COVID-19 infection; when *ϕ*_*R*_ = 1, no protection is present. The mosquito population consists of susceptible mosquitoes (*X*) and infected mosquitoes (*Y*). Recruitment of new mosquitoes occurs at a rate Ψ_*p*_, and all mosquitoes die naturally at a rate *µ*_*p*_. Susceptible mosquitoes become infected after biting infectious or partially infectious humans at a force of infection *λ*_*p*_, entering the infected class (*Y*). Infected mosquitoes transmit malaria to susceptible humans at a rate *λ*_*m*_.

The forces of infection governing transmission are defined as follows. The COVID-19 force of infection is

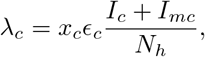

where *x*_*c*_*ϵ*_*c*_ represents effective human-to-human transmission. The malaria force of infection for humans is

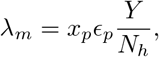

representing transmission from infected mosquitoes. The mosquito force of infection incorporates contributions from all malaria-infectious human classes,

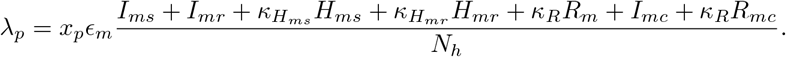

The parameters 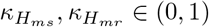 represent reduced infectiousness of treated malaria cases to mosquitoes, while *κ*_*R*_ ∈ (0, 1) represents reduced malaria transmissibility from recovered individuals with partial immunity. For simplicity, the same reduction factor *κ*_*R*_ is applied to both *R*_*m*_ and *R*_*mc*_. The resulting equation is presented below

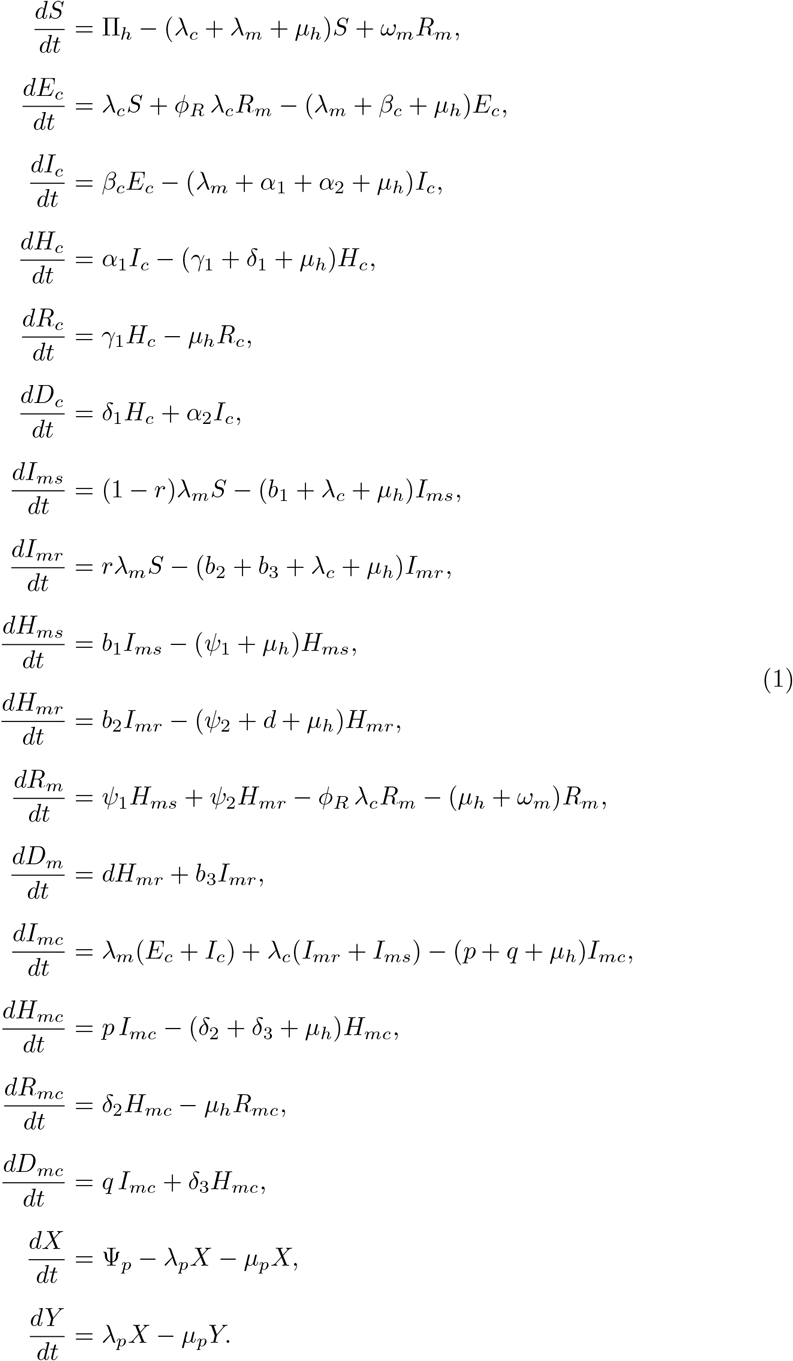

## Model Analysis

### Positivity and Boundedness of the Model solution

#### Lemma 1.

Assuming that

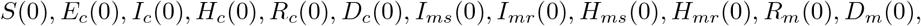

I_*mc*_(0), *H*_*mc*_(0), *R*_*mc*_(0), *D*_*mc*_(0), *X*(0), *Y* (0) ≥ 0,the corresponding solution remains nonnegative for all *t* ≥ 0.

#### Proposition 1.

For model (1) with nonnegative initial conditions, let

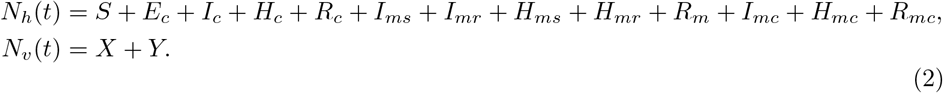

denote the total living human and vector populations, respectively. We define the feasible region

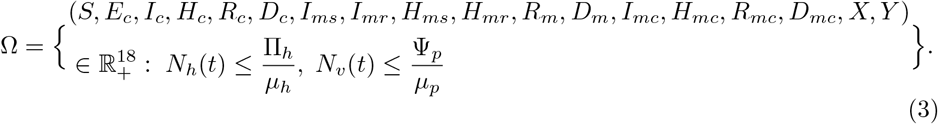

Under the flow of model (1), Ω is positively invariant and attracting.

*Proof*. Summing the equations of all *living* human compartments gives

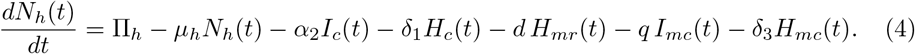

Here we have used the fact that all transfer terms between compartments (including the additional terms +*ω*_*m*_*R*_*m*_, −*ω*_*m*_*R*_*m*_ and ±*ϕ*_*R*_*λ*_*c*_*R*_*m*_) cancel in the sum. Similarly, adding the vector equations yields

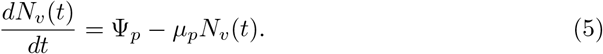

Hence,

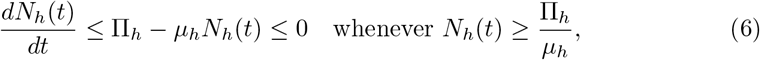

and

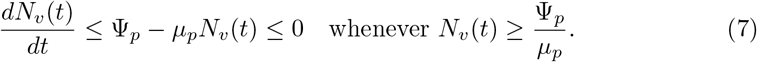

By comparison,

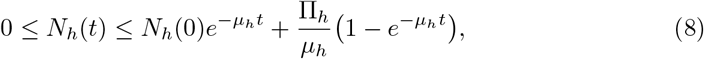

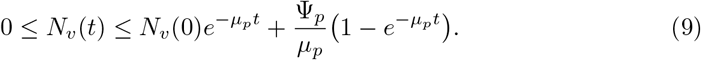

Thus, as *t* → ∞,

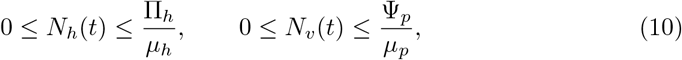

showing that Ω is attracting and positively invariant.

### Basic Reproduction Number

#### Derivation of ℛ_0,*c*_ (COVID only)

With malaria absent (*λ*_*m*_ ≡ 0) and all malaria compartments (including *R*_*m*_) identically zero, and assuming hospitalized individuals are non–infectious, the COVID sub–system is

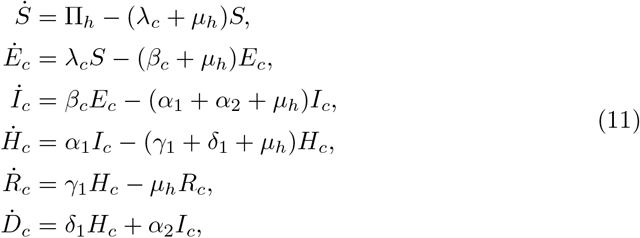

with force of infection

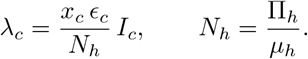

At the DFE, 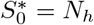 and all infected compartments vanish.

Now let **x** = (*E*_*c*_, *I*_*c*_, *H*_*c*_)^⊤^ and write 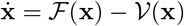, where

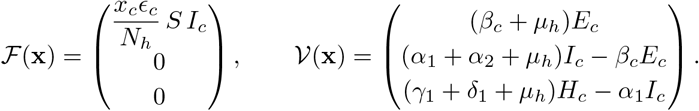

Then, the Jacobian matrices at the DFE are given by:

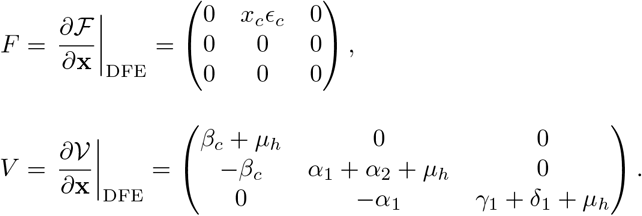

Then the inverse of *V* is:

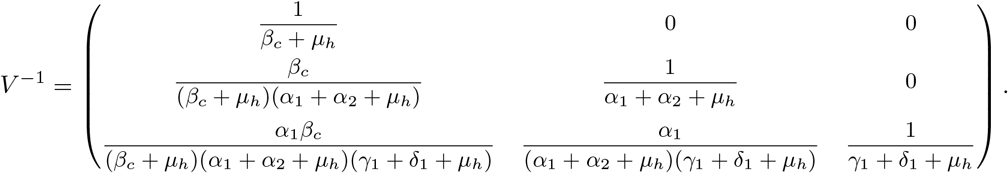

Therefore, this results in the next generation matrix:

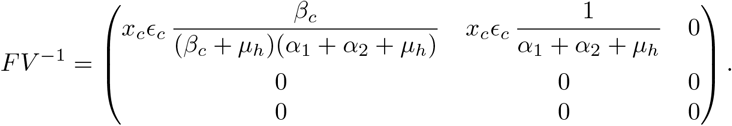

This matrix is upper–triangular, so its eigenvalues are its diagonal entries. The spectral radius is therefore

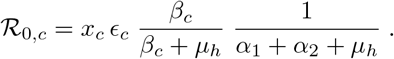

#### Derivation of ℛ_0,*m*_ (Malaria only)

With COVID–19 absent (*λ*_*c*_ ≡ 0) and all COVID compartments identically zero, the malaria sub–system reduces to

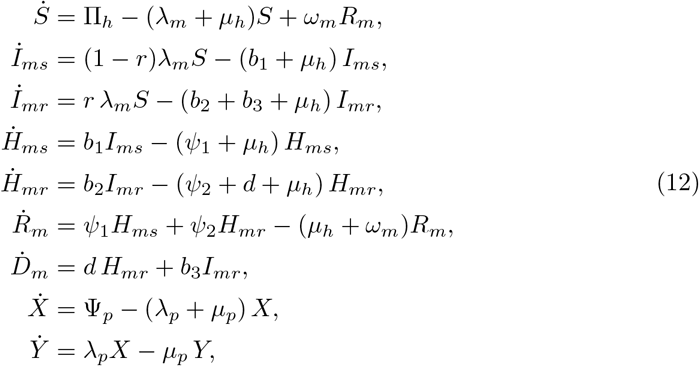

with forces of infection

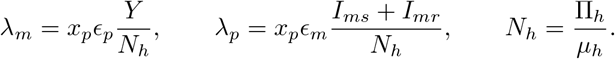

At the disease–free equilibrium (DFE),

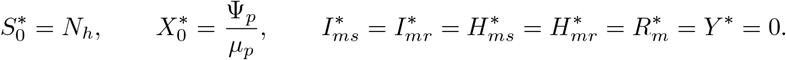

Note that the waning term *ω*_*m*_ does not affect the DFE 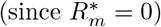 and does not enter the new infection terms.

We select the infected state vector

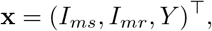

and decompose the system as 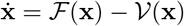, where

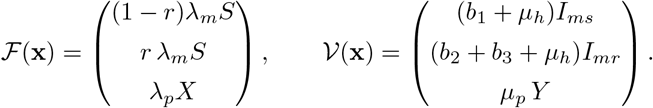

Evaluating the Jacobians at the DFE yields

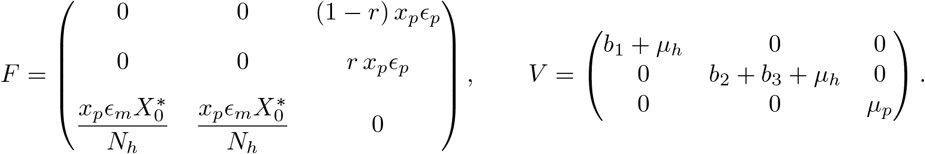

Since *V* is diagonal, its inverse is

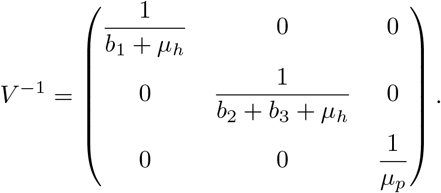

Thus, the next–generation matrix is

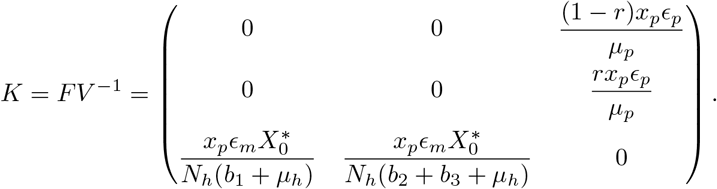

The basic reproduction number ℛ_0,*m*_ is given by the spectral radius of *K*, which yields

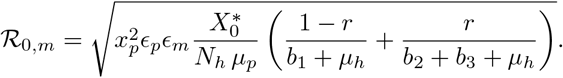

Introducing the mosquito–human ratio

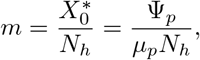

the malaria basic reproduction number can be written as

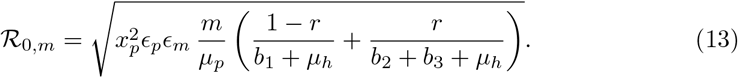

## Stochastic Model Formulation

Deterministic epidemic models assume smooth and predictable epidemic trajectories; however, real-world disease transmission is influenced by environmental variability, changes in mosquito abundance, fluctuations in human behaviour, and other sources of randomness. To account for these uncertainties in the co-dynamics of COVID–19 and malaria, we extend the deterministic model to a stochastic framework by introducing white-noise perturbations into the major transmission pathways. This produces a system of Itô-type stochastic differential equations (SDEs) which better reflect the variability observed in epidemiological data.

Let

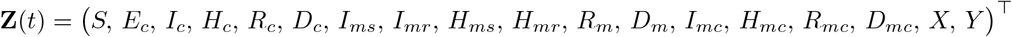

denote the state vector of the human and mosquito compartments. We introduce three independent standard Brownian motions,

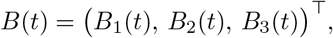

with intensities *σ*_1_ *>* 0, *σ*_2_ *>* 0, and *σ*_3_ *>* 0, representing random fluctuations in COVID transmission, malaria transmission to humans, and malaria transmission to mosquitoes, respectively. In the stochastic experiments reported here, we set

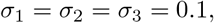

which corresponds to a common baseline noise level across the three transmission pathways. These values were not estimated from data, but were chosen to represent moderate stochastic variability and to illustrate the qualitative effect of transmission noise on the coupled dynamics.

The forces of infection retain their original deterministic structure,

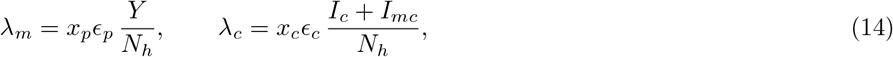

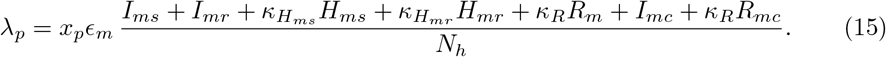

With stochastic perturbations introduced into the transmission terms, the complete stochastic COVID–19–malaria co-infection model takes the form

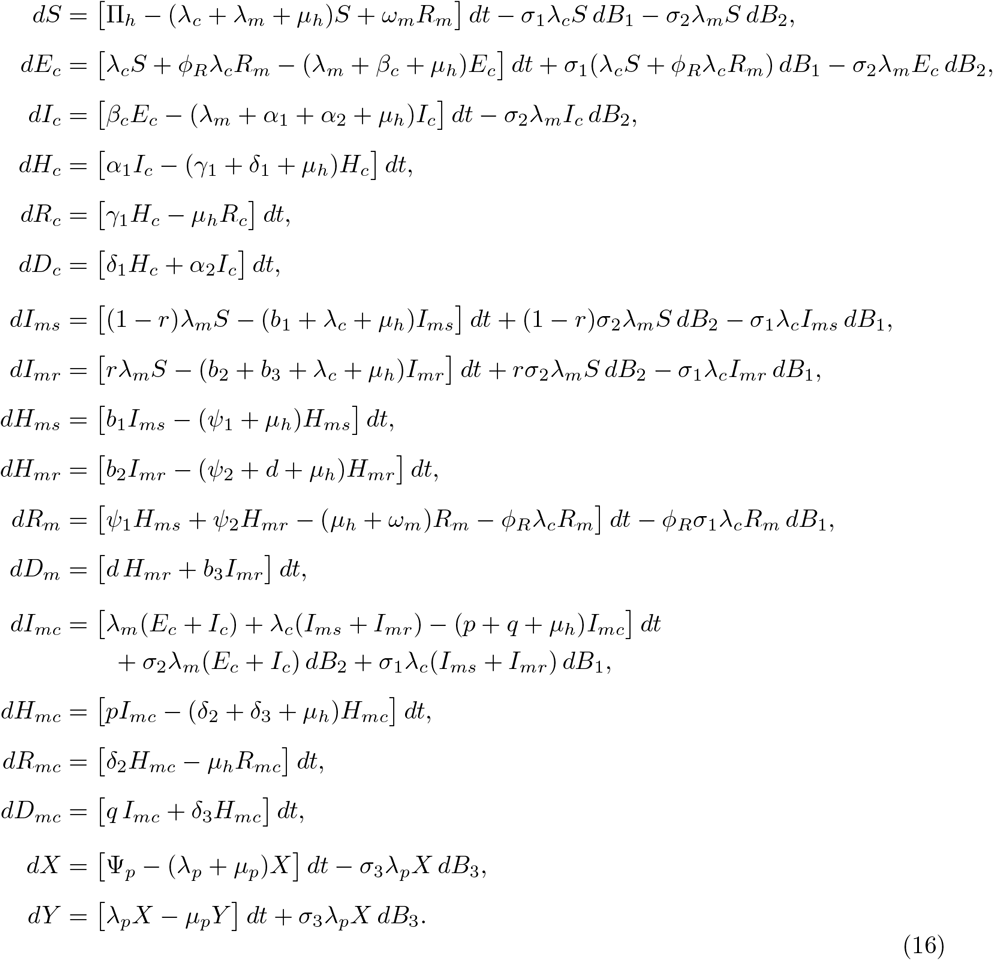

Equation (16) represents the full stochastic co-infection model, where the drift terms reproduce the deterministic dynamics and the diffusion terms produces random fluctuations.

### Remarks on stochastic well-posedness and simulation

The drift and diffusion coefficients in system (16) are continuously differentiable functions of the state variables and are therefore locally Lipschitz on the biologically relevant region. Consequently, for any nonnegative initial condition, the stochastic system admits a unique local strong solution up to a possible explosion time.

For numerical simulation of system (16), we employed the Euler–Maruyama scheme with time step Δ*t* = 0.1. For each scenario, 200 independent sample paths were generated from the prescribed initial conditions. The mean stochastic trajectory was computed pointwise across realizations, and the associated uncertainty band was obtained from the empirical 2.5% and 97.5% quantiles. No negative compartment values were observed in the simulations reported here; accordingly, no truncation, projection, or reflecting correction was applied.

A complete analytical proof of global positivity and boundedness for the full stochastic co-infection system is not provided here and is left for future study.

## Numerical Simulations

### Model Fitting

To parameterize the COVID-19 component of the proposed co-infection framework, we calibrated the COVID-only subsystem using Nigeria surveillance data for the early epidemic period, February 29 to June 27, 2020. Calibration was based on the cumulative numbers of confirmed cases and deaths reported in the Our World in Data (OWID) dataset. We used cumulative targets because daily incidence reports are often affected by reporting delays, weekend effects, and retrospective revisions, which can obscure the underlying epidemic trend. The fitting procedure used nonlinear least squares to estimate the effective transmission intensity of COVID-19. The remaining clinical progression parameters, including latent, hospitalization, recovery, and disease-induced mortality rates, were fixed at baseline values obtained from the literature (Table 1).

**Table 1.** Descriptions and baseline values for model parameters.

| Symbol | Description | Baseline value | Range | Source |
| --- | --- | --- | --- | --- |
| $\mu_h$ | Natural death rate of humans | $3.34 \times 10^{-5} \text{ day}^{-1}$ | – | [23] |
| $\Pi_h$ | Human recruitment (birth/immigration) (persons/day) | $\mu_h N_h$ | – | Derived |
| $\epsilon_c$ | Probability of infection per person-to-person contact | 0.01922 | (0, 1) | [22] |
| $x_c$ | Daily contact frequency per individual | $15.27 \text{ day}^{-1}$ | – | [24] |
| $\beta_c$ | Progression rate from exposed to infectious | $(1/5.1) \text{ day}^{-1}$ | – | [25] |
| $\alpha_1$ | Hospitalization rate for infectious COVID-19 cases ($I_c \rightarrow H_c$) | $0.154 \text{ day}^{-1}$ | [0.069, 0.183] | [26] |
| $\alpha_2$ | Disease-induced death rate for infectious COVID-19 individuals ($I_c \rightarrow D_c$) | $0.039925 \text{ day}^{-1}$ | – | [22] |
| $\gamma_1$ | Recovery rate for hospitalized COVID-19 cases ($H_c \rightarrow R_c$) | $0.16184 \text{ day}^{-1}$ | – | [22] |
| $\delta_1$ | Disease-induced death rate for hospitalized COVID-19 cases ($H_c \rightarrow D_c$) | $4.3665 \times 10^{-5} \text{ day}^{-1}$ | – | [22] |
| $\epsilon_p$ | Infection probability per mosquito-to-human bite | 0.48 | (0, 1) | [27] |
| $\epsilon_m$ | Infection probability per human-to-mosquito bite | 0.63896 | (0, 1) | [22] |
| $x_p$ | Average mosquito biting rate on humans | $4.3 \text{ day}^{-1}$ | (0.1, 50) | [27, 28] |
| $r$ | Proportion of new malaria infections that are drug-resistant | 0.3 | [0, 1] | [29, 30] |
| $b_1$ | Treatment initiation rate for drug-sensitive malaria ($I_{ms} \rightarrow H_{ms}$) | $0.33685 \text{ day}^{-1}$ | [0, 1] | [22] |
| $b_2$ | Treatment initiation rate for drug-resistant malaria ($I_{mr} \rightarrow H_{mr}$) | $0.33685 \text{ day}^{-1}$ | [0, 1] | [22] |
| $b_3$ | Malaria-induced death rate in drug-resistant infectious class | $0.001 \text{ day}^{-1}$ | [0, 0.005] | [31] |
| $d$ | Malaria-induced death rate in hospitalized malaria class | $0.01 \text{ day}^{-1}$ | [0, 0.05] | [31] |
| $\psi_1$ | Recovery rate for treated drug-sensitive malaria ($H_{ms} \rightarrow R_m$) | $0.10 \text{ day}^{-1}$ | [0.07, 0.14] | [29, 30] |
| $\psi_2$ | Recovery rate for treated drug-resistant malaria ($H_{mr} \rightarrow R_m$) | $0.07 \text{ day}^{-1}$ | [0.05, 0.10] | [29, 30] |
| $\omega_m$ | Waning rate of malaria-acquired immunity ($R_m \rightarrow S$) | $(1/365) \text{ day}^{-1}$ | [1/1825, 1/180] | [32] |
| $\kappa_{H_{ms}}$ | Relative infectiousness to mosquitoes of treated sensitive malaria cases | 0.5 | (0, 1) | Assumed |
| $\kappa_{H_{mr}}$ | Relative infectiousness to mosquitoes of treated resistant malaria cases | 0.5 | (0, 1) | Assumed |
| $\kappa_R$ | Relative infectiousness to mosquitoes of recovered/partially immune individuals | 0.5 | (0, 1) | Assumed |
| $\phi_R$ | Relative susceptibility of malaria-recovered individuals to COVID-19 ($\phi_R = 0$ full protection; $\phi_R = 1$ no protection) | Varied | [0, 1] | |
| $p$ | Hospitalization rate for co-infected individuals ($I_{mc} \rightarrow H_{mc}$) | $0.154 \text{ day}^{-1}$ | [0.069, 0.183] | [26] |
| $q$ | Disease-induced death rate for co-infected infectious individuals ($I_{mc} \rightarrow D_{mc}$) | $0.00629177 \text{ day}^{-1}$ | [0, 1] | [22] |
| $\delta_2$ | Recovery rate for hospitalized co-infected individuals ($H_{mc} \rightarrow R_{mc}$) | $0.0025851 \text{ day}^{-1}$ | [0, 1] | [22] |
| $\delta_3$ | Disease-induced death rate for hospitalized co-infected individuals ($H_{mc} \rightarrow D_{mc}$) | 0.001 | [0, 1] | [26] |
| $\Psi_p$ | Mosquito recruitment rate | $1000 \text{ day}^{-1}$ | – | [33] |
| $\mu_p$ | Mosquito natural death rate | $0.033 \text{ day}^{-1}$ | [0.01, 0.1] | [28] |

For malaria, because daily malaria surveillance time-series were not available for direct fitting, the malaria-only subsystem was used to construct an endemic baseline consistent with reported annual malaria cases and deaths in Nigeria for 2021. Most malaria parameters were fixed from the literature, while the mosquito-to-human ratio and an overall malaria transmission scaling parameter were assigned baseline values and varied under uncertainty analysis. To obtain realistic endemic dynamics, the malaria subsystem was simulated with seasonal mosquito recruitment and a burn-in period before extracting a one-year observation window. The resulting annual malaria burden was then compared with the target annual case and death totals, and uncertainty bands were generated from repeated simulations under parameter variation. Consequently, the malaria component should be interpreted as an endemic baseline consistent with annual burden estimates rather than as a direct fit to daily malaria observations. For the full COVID-19-malaria co-infection model, parameters were therefore classified as data-estimated, literature-based, or specified for scenario exploration (Table 1). In particular, co-infection interaction parameters, including those related to malaria-acquired partial protection, were not directly estimated from joint COVID-19–malaria data, but assigned biologically plausible baseline values and examined through numerical simulations and sensitivity analysis. Thus, the full coupled model is intended primarily as a mechanistic and scenario-based framework rather than a fully data-calibrated forecasting model.

Fig 3(a) shows the model fit to cumulative cases, whereas Fig 3(b) shows the fit to cumulative deaths. The fitted model reproduces the overall growth pattern and curvature of both cumulative time series over the calibration period, capturing the accelerating increase during the latter portion of the window and remaining close to the observed trajectories throughout. Small deviations between model and data are expected, particularly because cumulative reported cases depend on evolving testing capacity and reporting practices. In contrast, cumulative deaths may be affected by delays in notification and under-ascertainment. however, the close agreement indicates that the calibrated COVID-only subsystem provides a reasonable quantitative description of early COVID-19 transmission and mortality dynamics in Nigeria within a parsimonious constant-parameter framework.

**Fig 3.**
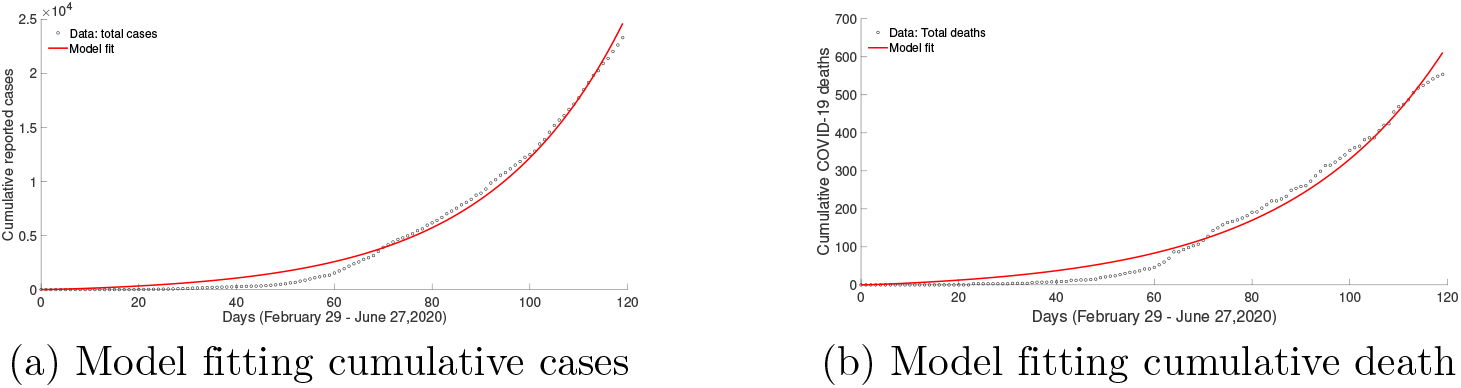
Model fitting results for cumulative cases and deaths. (Data source: Our World in Data [34]).

The malaria calibration results in Fig 4 summarize the modeled daily malaria burden under parameter uncertainty. Fig 4(a) shows the modeled daily malaria incidence together with the 95% uncertainty interval band (2.5th–97.5th percentiles), while Fig 4(b) presents the corresponding daily malaria deaths. The trajectories display smooth endemic dynamics consistent with sustained malaria transmission, and the uncertainty bands quantify variability arising from uncertainty in the fitted transmission intensity and mosquito-to-human ratio. Importantly, the modeled daily profiles are consistent with the annual malaria case and death totals used for calibration, providing a realistic endemic baseline for subsequent simulations of the coupled COVID–19–malaria co-infection model.

**Fig 4.**
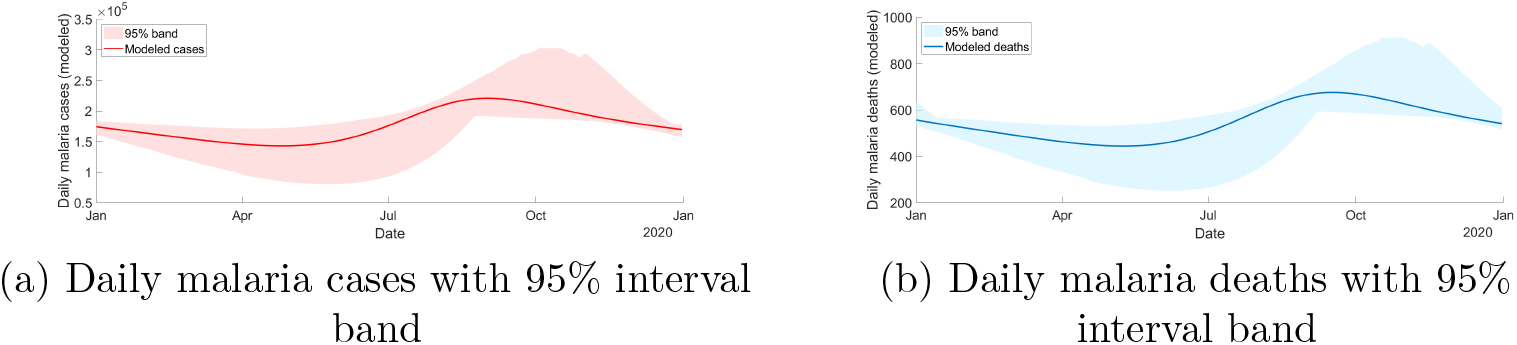
Modeled daily malaria cases and deaths with 95% uncertainty interval band (2.5th-97.5th percentiles)

### Results and Interpretation

Fig 5(a) compares the deterministic trajectory with the stochastic dynamics of the combined COVID-19 infectious burden, *I*_*c*_(*t*) + *I*_*mc*_(*t*), obtained from multiple stochastic realizations. The grey shaded region represents the 95% confidence band, the solid red curve denotes the stochastic mean, and the dashed blue curve corresponds to the deterministic solution. The deterministic trajectory remains entirely within the stochastic confidence envelope, indicating that the mean-field dynamics are broadly consistent with the stochastic formulation. Nevertheless, noticeable differences arise in both the timing and magnitude of the epidemic peak, with stochastic fluctuations producing an earlier and higher peak relative to the deterministic prediction. Such behavior is expected in nonlinear epidemic systems driven by multiplicative Itô noise, for which the ensemble mean of the stochastic process does not necessarily coincide with the deterministic solution. The widening of the confidence band at later times reflects sustained variability induced by random perturbations in transmission processes.

**Fig 5.**
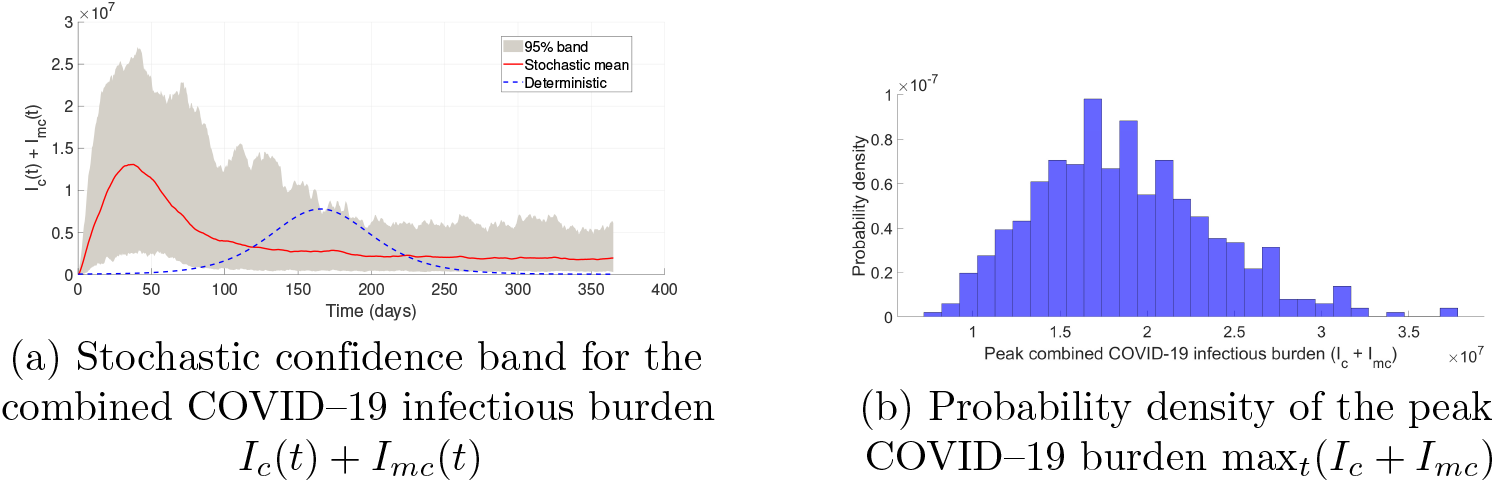
Stochastic analysis of COVID–19 dynamics under random transmission effects. Panel (a) shows the deterministic trajectory together with the stochastic mean and the 95% confidence band for the combined COVID–19 infectious burden. Panel (b) presents the probability distribution of the corresponding epidemic peak obtained from multiple stochastic realizations, illustrating the variability and risk of large outbreak events.

Fig 5(b) presents the probability distribution of the peak combined COVID–19 infectious burden, max_*t*_(*I*_*c*_ + *I*_*mc*_), across stochastic simulations. While most realizations cluster around a typical epidemic peak, the distribution exhibits substantial variability and a pronounced right-skewed tail, indicating that large outbreaks, although less frequent, remain possible and are not captured by deterministic predictions alone. Together, these results highlight the importance of incorporating stochastic effects when assessing epidemic uncertainty and outbreak risk in malaria-endemic settings.

### Sensitivity Analysis Using Partial Rank Correlation Coefficients

A global sensitivity analysis was conducted using *Partial Rank Correlation Coefficients* (PRCCs) to identify parameters that most strongly influence COVID–19 dynamics in the presence of malaria co-infection. PRCCs quantify the strength and direction of monotonic relationships between uncertain model parameters and selected epidemiological outcomes while controlling for the effects of all other parameters. This method is well suited for nonlinear epidemic models and has been widely applied in mathematical epidemiology.

The sensitivity analysis was performed on epidemiologically relevant model outputs, rather than reproduction numbers, which are defined at the disease-free equilibrium and do not capture the effects of malaria-acquired immunity or co-infection dynamics. Two COVID–19 outcomes were considered: (i) the *peak COVID burden*, defined as

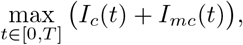

and (ii) the *cumulative COVID–19 mortality*, measured by the final number of COVID–19 deaths *D*_*c*_(*T*) at the end of the simulation period. Key COVID–19, malaria, co-infection, and mosquito-related parameters were varied simultaneously over biologically plausible ranges using Latin Hypercube Sampling. For each sampled parameter set, the deterministic model was solved numerically over a one-year simulation horizon, and the corresponding outcome measures were recorded. Statistical significance of PRCC values was assessed at the 5% level.

Fig 6(a) presents the PRCC results for the peak COVID burden max(*I*_*c*_ + *I*_*mc*_). The effective contact rate of COVID-19 *x*_*c*_ and the transmission probability per contact *ϵ*_*c*_ exhibit strong positive PRCC values, indicating that the increase in transmission intensity substantially amplifies the maximum COVID burden. In contrast, the hospitalization rate *α*_1_ displays a strong negative PRCC, reflecting the reduction in infectious prevalence associated with faster removal of infectious individuals through hospitalization. Other COVID–19 progression parameters contribute to the peak burden with comparatively smaller magnitudes.

**Fig 6.**
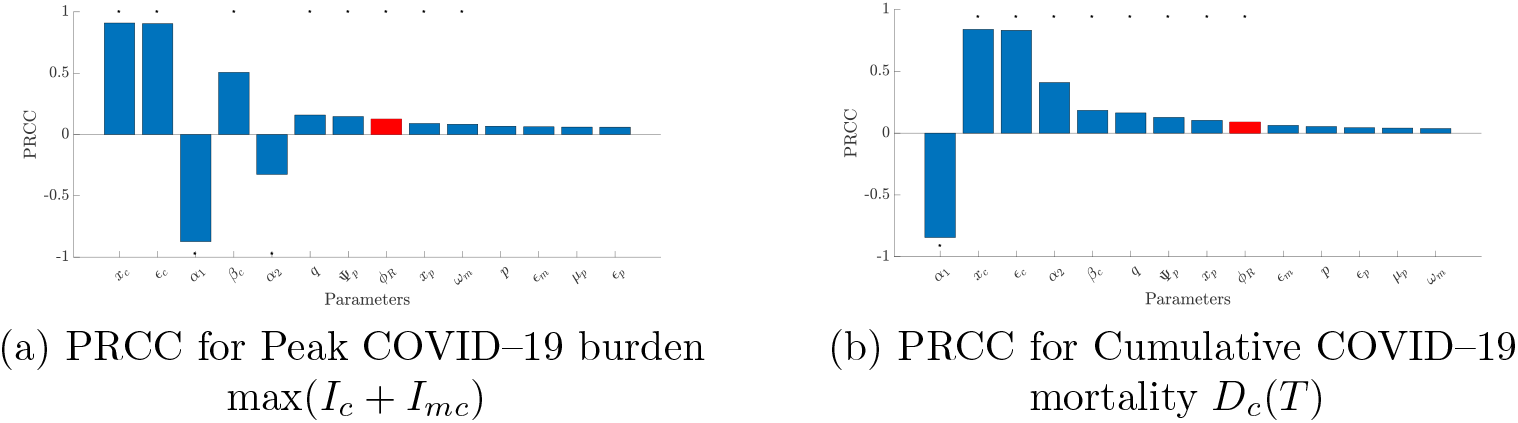
Partial rank correlation coefficients (PRCCs) for COVID-19 outcomes. Panel (a) shows PRCC values for the peak COVID–19 burden max(*I*_*c*_ + *I*_*mc*_), while panel (b) presents PRCC results for cumulative COVID-19 mortality *D*_*c*_(*T*). Bars represent the strength and direction of the monotonic relationship between model parameters and the corresponding outcome after controlling for the effects of all other parameters.

The malaria-immunity parameter *ϕ*_*R*_ shows a *positive and statistically significant* PRCC with the maximum burden of COVID. Since *ϕ*_*R*_ represents the relative susceptibility of malaria-recovered individuals to COVID–19 infection, this result indicates that reduced malaria-derived protection (larger *ϕ*_*R*_) is associated with higher peak COVID prevalence. Conversely, stronger malaria-acquired immunity contributes to a reduction in the COVID-19 epidemic peak. Although the magnitude of the PRCC associated with *ϕ*_*R*_ is smaller than that of the primary COVID-19 transmission parameters, its statistical significance highlights the non-negligible role of malaria-induced immunity in shaping COVID-19 dynamics.

Fig 6(b) shows the PRCC results for cumulative mortality from COVID–19 *D*_*c*_(*T*). Similar trends are observed, with the transmission parameters of COVID-19 exerting the strongest influence on mortality outcomes, while the hospitalization rate *α*_1_ remains strongly negatively correlated with cumulative deaths. Importantly, the parameter *ϕ*_*R*_ again shows a *positive and statistically significant* PRCC, indicating that weaker malaria-acquired immunity leads to increased mortality from COVID-19. This finding suggests that malaria-induced immune effects may indirectly reduce COVID-19 related deaths by lowering susceptibility and subsequent disease progression among malaria-recovered individuals.

To further complement the PRCC results and visualize the interaction effects between the influential parameters, three-dimensional surface plots were constructed to examine the combined impact of the malaria-acquired immunity parameter *ϕ*_*R*_ and the COVID-19 contact rate *x*_*c*_ on key epidemiological outcomes. As shown in Figs 7(a) and 7(b), increasing *x*_*c*_ leads to a pronounced increase in both the peak COVID-19 burden, measured by max(*I*_*c*_ + *I*_*mc*_), and the cumulative mortality from COVID-19 *D*_*c*_(*T*), confirming the dominant role of transmission intensity identified by the PRCC analysis. In contrast, lower values of *ϕ*_*R*_ consistently reduce both outcomes at all levels of *x*_*c*_, reflecting the mitigating effect of malaria-acquired immunity on COVID-19 susceptibility among malaria-recovered individuals. In particular, the protective influence of *ϕ*_*R*_ becomes more evident at intermediate and high contact rates, indicating that malaria-induced immunity can partially offset the severity of COVID-19 in high-transmission settings.

**Fig 7.**
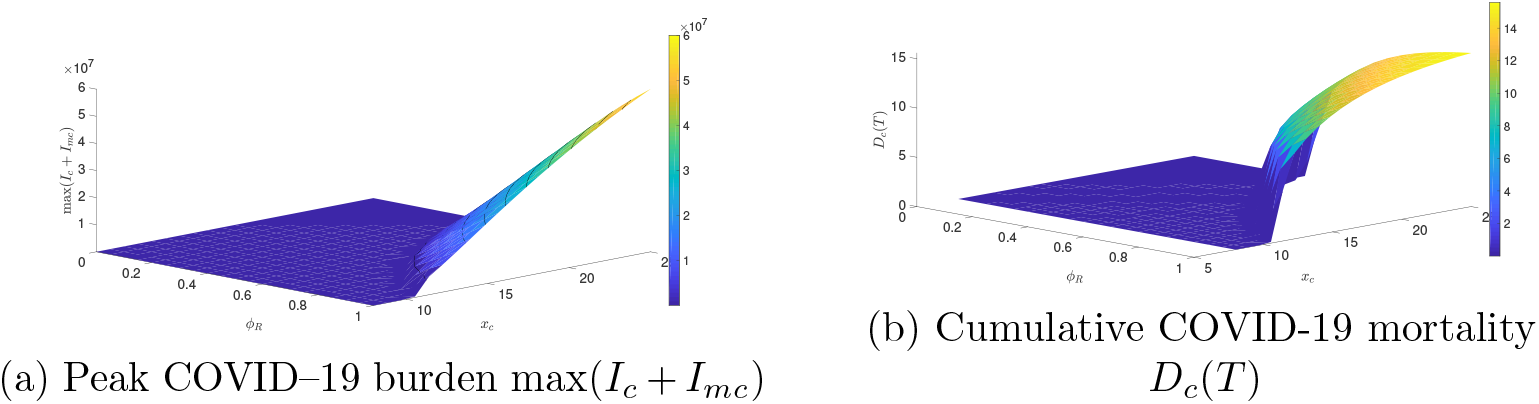
Three-dimensional surface plots illustrating the combined effects of malaria–acquired immunity and COVID–19 transmission intensity on COVID–19 outcomes. Panel (a) shows the variation in the peak COVID-19 burden max(*I*_*c*_ + *I*_*mc*_) as a function of the malaria immunity parameter *ϕ*_*R*_ and the COVID–19 contact rate *x*_*c*_. Panel (b) depicts the corresponding effects on cumulative COVID-19 mortality *D*_*c*_(*T*).

## Conclusion

In this study, we developed a deterministic-stochastic compartmental model for the joint transmission of COVID-19 and malaria in endemic settings. The framework incorporates key epidemiological features of both diseases, including mosquito-borne malaria transmission, hospitalization, disease-induced mortality, co-infection, waning malaria immunity, and the presence of both drug-sensitive and drug-resistant malaria strains. A particularly important aspect of the model is the inclusion of malaria-acquired partial immunity through the parameter *ϕ*_*R*_, which was used to represent reduced susceptibility to COVID-19 among individuals who had recovered from malaria.

On the analytical side, we showed that the deterministic system is mathematically well posed by establishing positivity, boundedness, and the existence of an invariant feasible region. We also derived the basic reproduction numbers for the COVID-19-only and malaria-only subsystems, which provide useful threshold quantities for understanding transmission in the absence of the other disease.

The numerical results indicate that, within the assumptions of the model, malaria-acquired partial immunity may help reduce the burden of COVID-19 by lowering the peak number of infectious individuals, reducing hospitalization pressure, and decreasing cumulative mortality. At the same time, the results also suggest that strong malaria transmission and the persistence of drug resistance can continue to sustain malaria burden and increase the possibility of co-infection. Taken together, these findings show that the interaction between the two diseases may matter in important ways in endemic regions.

The stochastic version of the model adds another layer to this picture by showing that random variation in transmission can lead to noticeable differences in epidemic behavior. In particular, the stochastic simulations showed variation in both the timing and the size of outbreak peaks, and they also pointed to the possibility of larger outbreaks than those predicted by the deterministic model alone. This highlights the importance of accounting for uncertainty when studying co-infection dynamics, especially in settings where environmental and behavioral conditions can vary over time.

The sensitivity analysis further showed that the main COVID-19 transmission parameters had the strongest influence on peak burden and mortality, while increased hospitalization tended to reduce both outcomes. The parameter *ϕ*_*R*_ was also found to have a statistically significant effect on these outcomes, suggesting that malaria-related immune effects may play a meaningful role in shaping COVID-19 dynamics within the modeling framework considered here.

Overall, this work provides a mechanistic framework for studying the coupled dynamics of COVID-19 and malaria while also accounting for uncertainty in transmission. Since the malaria and co-infection components were not fully calibrated to joint time-series data, the results should be viewed mainly as scenario-based and exploratory rather than as direct forecasts. Future work could build on this study by improving the empirical estimation of immunity-related effects, incorporating intervention measures and seasonal variation more fully, and calibrating the complete co-infection system to richer joint datasets. These extensions would help strengthen the practical relevance of the model for understanding overlapping respiratory and vector-borne disease risks in endemic settings.

## Supporting information

Latex

## Data Availability

All data produced in the present work are contained in the manuscript

## Acknowledgment

We would like to thank the support of National Science Foundation (DMS-2533878, DMS-2053746, DMS-2134209, ECCS-2328241, CBET-2347401 and OAC-2311848), and U.S. Department of Energy (DOE) Office of Science Advanced Scientific Computing Research program DE-SC0023161, the SciDAC LEADS Institute, and DOE–Fusion Energy Science, under grant number: DE-SC0024583.

