## Supplementary material for "A Deterministic–Stochastic Model for COVID-19 and Malaria Co-Infection with Malaria-Acquired Partial Immunity": Latex: makefile

```
# $Id: makefile 193 2020-11-23 11:15:37Z rishi $

file=elsdoc


all: pdf out 
	make pdf
	make pdf

out:
	if  [ -f $(file).out ] ; then cp $(file).out tmp.out; fi ;
	sed 's/BOOKMARK/dtxmark/g;' tmp.out > x.out; mv x.out tmp.out ;

pdf:
	pdflatex $(file).tex

index:
	makeindex -s gind.ist -o $(file).ind $(file).idx 

changes:
	makeindex -s gglo.ist -o $(file).gls $(file).glo

xview:
#	xpdf -z 200 $(file).pdf &>/dev/null
	open -a 'Skim.app' $(file).pdf 

view:
	open -a 'Adobe Reader.app' $(file).pdf

ins:
	latex $(file).ins 

diff:
	diff $(file).sty ../$(file).sty |less

copy:
	cp $(file).sty ../
```
