## Supplementary material for "A Deterministic–Stochastic Model for COVID-19 and Malaria Co-Infection with Malaria-Acquired Partial Immunity": Latex: manifest.txt

% Copyright 2007-2020 Elsevier Ltd
%
% This file is part of the 'Elsarticle Bundle'.
% ---------------------------------------------
%
% It may be distributed and/or modified under the
% conditions of the LaTeX Project Public License, either version 1.2
% of this license or (at your option) any later version.
% The latest version of this license is in
%    http://www.latex-project.org/lppl.txt
% and version 1.2 or later is part of all distributions of LaTeX
% version 1999/12/01 or later.
%
% The list of all files belonging to the LaTeX 'Elsarticle Bundle' is
% given in the file `manifest.txt'.
%
% CONTENTS OF THE ELSARTICLE BUNDLE
% ===============================

Directory elsarticle/

elsarticle.dtx
        Main package with driver

elsarticle.ins
  installer for elsarticle.dtx

elsarticle-harv.bst
  bibliographic style for author-year citation

elsarticle-num.bst
  bibliographic style for numbered citation

elsarticle-num-names.bst
  bibliographic style for numbered citation + new natbib options

elsarticle-template-harv.tex
  article template for a document with author-year citation
 
elsarticle-template-num.tex
  article template for a document with numbered citation

manifest.txt
  this file

README
  small readme documentation

Directory elsarticle/doc/

The following files are graphic files needed for creating pdf output
of the documentation from elsdoc.tex:

 1pseperateaug.pdf
 1psingleauthorgroup.pdf
 elstest-1pdoubleblind.pdf
 elstest-1p.pdf
 elstest-3pd.pdf
 elstest-3p.pdf
 elstest-5p.pdf
 jfigs.pdf

elsdoc.tex  -- LaTeX source file of documentation
elsdoc.pdf  -- documentation for elsarticle.cls

The following files are files written out every time elsdoc.tex is
compiled:

 elsdoc.aux
 elsdoc.log
 elsdoc.out
 tmp.tex

Auxiliary packages needed to generate pdf output from elsdoc.tex:

 rvdtx.sty
 pdfwidgets.sty
