## Supplementary material for "A Deterministic–Stochastic Model for COVID-19 and Malaria Co-Infection with Malaria-Acquired Partial Immunity": Latex: plos_bibtex_sample.bib

@article{avusuglo2024assessment,
  title={Assessment of bidirectional impact of stigmatization induced self-medication on COVID-19 and malaria transmissions using mathematical modeling: Nigeria as a case study},
  author={Avusuglo, Wisdom S and Han, Qing and Woldegerima, Woldegebriel Assefa and Asgary, Ali and Wu, Jianhong and Orbinski, James and Bragazzi, Nicola and Ahmadi, Ali and Kong, Jude Dzevela},
  journal={Mathematical Biosciences},
  volume={376},
  pages={109249},
  year={2024},
  doi = {https://doi.org/10.1016/j.mbs.2024.109249}
  publisher={Elsevier}
}

@article{li2024analysis,
  title={Analysis of a delayed malaria transmission model including vaccination with waning immunity and reinfection},
  author={Li, Jinhui and Teng, Zhidong and Wang, Ning and Chen, Wei},
  journal={Journal of Applied Mathematics and Computing},
  volume={70},
  number={4},
  pages={3917--3946},
  year={2024},
  doi = {https://doi.org/10.1007/s12190-024-02124-1}
  publisher={Springer}
}

@article{chitnis2008determining,
  title={Determining important parameters in the spread of malaria through the sensitivity analysis of a mathematical model},
  author={Chitnis, Nakul and Hyman, James M and Cushing, Jim M},
  journal={Bulletin of mathematical biology},
  volume={70},
  number={5},
  pages={1272--1296},
  year={2008},
  doi = {https://doi.org/10.1007/s11538-008-9299-0}
  publisher={Springer}
}

@article{oluwatobi2023impact,
  title={Impact of Contaminated Surfaces on the Transmission Dynamics of Corona Virus Disease (Covid-19)},
  author={Oluwatobi, Kabir},
  journal={Biomedical Journal of Scientific \& Technical Research},
  volume={51},
  number={1},
  pages={42280--42294},
  year={2023},
  doi = {https://doi.org/10.26717/BJSTR.2023.51.008046}
  publisher={Biomedical Research Network+, LLC}
}

@article{adeniyi2020dynamic,
  title={Dynamic model of COVID-19 disease with exploratory data analysis},
  author={Adeniyi, Michael O and Ekum, Matthew I and Oke, Segun I and others},
  journal={Scientific African},
  volume={9},
  pages={e00477},
  year={2020},
  doi = {https://doi.org/10.1016/j.sciaf.2020.e00477}
  publisher={Elsevier}
}

@article{prem2021projecting,
  title={Projecting contact matrices in 177 geographical regions: An update and comparison with empirical data for the COVID-19 era},
  author={Prem, Kiesha and Zandvoort, Kevin van and Klepac, Petra and Eggo, Rosalind M and Davies, Nicholas G and Centre for the Mathematical Modelling of Infectious Diseases COVID-19 Working Group and Cook, Alex R and Jit, Mark},
  journal={PLoS computational biology},
  volume={17},
  number={7},
  pages={e1009098},
  year={2021},
  doi = {https://doi.org/10.1371/journal.pcbi.1009098}
  publisher={Public Library of Science San Francisco, CA USA}
}

@article{lauer2020incubation,
  title={The incubation period of coronavirus disease 2019 (COVID-19) from publicly reported confirmed cases: estimation and application},
  author={Lauer, Stephen A and Grantz, Kyra H and Bi, Qifang and Jones, Forrest K and Zheng, Qulu and Meredith, Hannah R and Azman, Andrew S and Reich, Nicholas G and Lessler, Justin},
  journal={Annals of internal medicine},
  volume={172},
  number={9},
  pages={577--582},
  year={2020},
  doi = {https://doi.org/10.7326/M20-0504}
  publisher={American College of Physicians}
}

@article{iboi2020mathematical,
  title={Mathematical modeling and analysis of COVID-19 pandemic in Nigeria},
  author={Iboi, Enahoro and Sharomi, Oluwaseun O and Ngonghala, Calistus and Gumel, Abba B},
  journal={MedRxiv},
  pages={2020--05},
  year={2020},
  doi = {https://doi.org/10.1101/2020.05.22.20110387}
  publisher={Cold Spring Harbor Laboratory Press}
}

@article{herdicho2021optimal,
  title={An optimal control of malaria transmission model with mosquito seasonal factor},
  author={Herdicho, Faishal Farrel and Chukwu, Williams and Tasman, Hengki and others},
  journal={Results in physics},
  volume={25},
  pages={104238},
  year={2021},
  doi = {https://doi.org/10.1016/j.rinp.2021.104238}
  publisher={Elsevier}
}

@article{collins2022mathematical,
  title={A mathematical model for the dynamics and control of malaria in Nigeria},
  author={Collins, OC and Duffy, KJ},
  journal={Infectious disease modelling},
  volume={7},
  number={4},
  pages={728--741},
  year={2022},
  doi = {https://doi.org/10.1016/j.idm.2022.10.005}
  publisher={Elsevier}
}

@article{bakare2017bifurcation,
  title={Bifurcation and sensitivity analysis of malaria--schistosomiasis co-infection model},
  author={Bakare, EA and Nwozo, CR},
  journal={International Journal of Applied and Computational Mathematics},
  volume={3},
  pages={971--1000},
  year={2017},
  doi ={https://doi.org/10.1007/s40819-017-0394-5}
  publisher={Springer}
}

@article{chitnis2008determining,
  title={Determining important parameters in the spread of malaria through the sensitivity analysis of a mathematical model},
  author={Chitnis, Nakul and Hyman, James M and Cushing, Jim M},
  journal={Bulletin of mathematical biology},
  volume={70},
  pages={1272--1296},
  year={2008},
  doi = {https://doi.org/10.1007/s11538-008-9299-0}
  publisher={Springer}
}

@article{tumwiine2014mathematical,
  title={A mathematical model for the transmission and spread of drug sensitive and resistant malaria strains within a human population},
  author={Tumwiine, Julius and Hove-Musekwa, Senelani D and Nyabadza, Farai},
  journal={International Scholarly Research Notices},
  volume={2014},
  number={1},
  pages={636973},
  year={2014},
  doi ={ https://doi.org/10.1155/2014/636973}
  publisher={Wiley Online Library}
}

@article{gemperli2006malaria,
  title={Malaria mapping using transmission models: application to survey data from Mali},
  author={Gemperli, A and Vounatsou, P and Sogoba, N and Smith, T},
  journal={American journal of Epidemiology},
  volume={163},
  number={3},
  pages={289--297},
  year={2006},
  doi = { https://doi.org/10.1093/aje/kwj026}
  publisher={Oxford University Press}
}

@article{barnes2005population,
  title={Population biology and antimalarial resistance: The transmission of antimalarial drug resistance in Plasmodium falciparum},
  author={Barnes, Karen I and White, Nicholas J},
  journal={Acta tropica},
  volume={94},
  number={3},
  pages={230--240},
  year={2005},
  doi = {https://doi.org/10.1016/j.actatropica.2005.04.014}
  publisher={Elsevier}
}

@article{bala2019global,
  title={Global sensitivity analysis to study the impacts of bed-nets, drug treatment, and their efficacies on a two-strain malaria model},
  author={Bala, Saminu and Gimba, Bello},
  journal={Mathematical and Computational Applications},
  volume={24},
  number={1},
  pages={32},
  year={2019},
  doi = {https://doi.org/10.3390/mca24010032}
  publisher={MDPI}
}

@article{woldegerima2021mathematical,
  title={Mathematical analysis of the impact of transmission-blocking drugs on the population dynamics of malaria},
  author={Woldegerima, Woldegebriel Assefa and Ouifki, Rachid and Banasiak, Jacek},
  journal={Applied Mathematics and Computation},
  volume={400},
  pages={126005},
  year={2021},
  doi = {https://doi.org/10.1016/j.amc.2021.126005}
  publisher={Elsevier}
}

@article{park2023barriers,
  title={Barriers against and strategies for malaria control during the COVID-19 pandemic in low-and middle-income countries: a systematic review},
  author={Park, Jiwook and Kang, Seungwoo and Seok, Dayoung and Baek, Yae Jee and An, Se Young and Lee, Junga and Jun, Alina and Kim, Sun-Young},
  journal={Malaria journal},
  volume={22},
  number={1},
  pages={41},
  year={2023},
  doi = {https://doi.org/10.1186/s12936-023-04452-2}
  publisher={Springer}
}

@article{moytrayee2025maintaining,
  title={Maintaining essential health services during COVID-19: cross-country lessons of health system resilience from Asia, Sub-Saharan Africa and Latin America},
  author={Moytrayee, Guha and Cohen, Tanugi-Carresse Arielle and Mullen, Lucia and Bennett, Sara and Kitti, Wanyenze Rhoda and Prado, Andrea M and Piya, Hanvoravongchai and Magdalena, Rathe and Julius, Fobil and Prasan, Rannan-Eliya Ravindra and others},
  journal={BMJ Global Health},
  volume={8},
  number={Suppl 6},
  pages={e013392},
  year={2025},
  doi = {https://doi.org/10.1136/bmjgh-2023-013392}
  publisher={BMJ Publishing Group LTD}
}

@article{zulu2025scoping,
  title={A scoping review of the roles, challenges, and strategies for enhancing the performance of community health workers in the response against COVID-19 in low-and middle-income countries},
  author={Zulu, Joseph Mumba and Silumbwe, Adam and Munakampe, Margarate and Chavula, Malizgani Paul and Mulubwa, Chama and Sirili, Nathanael and Zulu, Wanga and Michelo, Charles and Tetui, Moses},
  journal={BMC Primary Care},
  volume={26},
  number={1},
  pages={163},
  year={2025},
  doi = {https://doi.org/10.1186/s12875-025-02853-7}
  publisher={Springer}
}

@article{kolawole2023malaria,
  title={Malaria endemicity in Sub-Saharan Africa: Past and present issues in public health},
  author={Kolawole, Eniola O and Ayeni, Eunice T and Abolade, Sodiq A and Ugwu, Stephen E and Awoyinka, Temitope B and Ofeh, Augustine S and Okolo, Bartholomew O},
  journal={Microbes and Infectious Diseases},
  volume={4},
  number={1},
  pages={242--251},
  year={2023},
  doi = {https://doi.org/10.21608/mid.2022.150194.1346}
  publisher={Zagazig University, Faculty of Medicine}
}

@article{konozy2022striking,
  title={The striking mimics between COVID-19 and malaria: A review},
  author={Konozy, Emadeldin Hassan E and Osman, Makarim El-fadil M and Ghartey-Kwansah, George and Abushama, Hind Mohamed},
  journal={Frontiers in Immunology},
  volume={13},
  pages={957913},
  year={2022},
  doi = {https://doi.org/10.3389/fimmu.2022.957913}
  publisher={Frontiers Media SA}
}

@article{cohen2022fight,
  title={The fight against malaria: diminishing gains and growing challenges},
  author={Cohen, Justin M and Okumu, Fredros and Moonen, Bruno},
  journal={Science Translational Medicine},
  volume={14},
  number={651},
  pages={eabn3256},
  year={2022},
  doi = {https://doi.org/10.1126/scitranslmed.abn3256}
  publisher={American Association for the Advancement of Science}
}

@article{siddique2021unprecedented,
  title={Unprecedented environmental and energy impacts and challenges of COVID-19 pandemic},
  author={Siddique, Azhar and Shahzad, Asif and Lawler, Jenny and Mahmoud, Khaled A and Lee, Dae Sung and Ali, Nisar and Bilal, Muhammad and Rasool, Kashif},
  journal={Environmental Research},
  volume={193},
  pages={110443},
  year={2021},
  doi = {https://doi.org/10.1016/j.envres.2020.110443}
  publisher={Elsevier}
}

@article{hussein2020malaria,
  title={Malaria and COVID-19: unmasking their ties},
  author={Hussein, Mogahed Ismail Hassan and Albashir, Ahmed Abdalazim Dafallah and Elawad, Omer Ali Mohamed Ahmed and Homeida, Anmar},
  journal={Malaria journal},
  volume={19},
  number={1},
  pages={457},
  year={2020},
  doi = {https://doi.org/10.1186/s12936-020-03541-w}
  publisher={Springer}
}

@article{hussein2022impact,
  title={Impact of COVID-19 and malaria coinfection on clinical outcomes: a retrospective cohort study},
  author={Hussein, Rasha and Guedes, Murilo and Ibraheim, Nada and Ali, Mazin M and El-Tahir, Amal and Allam, Nahla and Abuakar, Hussain and Pecoits-Filho, Roberto and Kotanko, Peter},
  journal={Clinical Microbiology and Infection},
  volume={28},
  number={8},
  pages={1152--e1},
  year={2022},
  doi = {https://doi.org/10.1016/j.cmi.2022.03.028}
  publisher={Elsevier}
}

@article{anyanwu2021association,
  title={The association between malaria prevalence and COVID-19 mortality},
  author={Anyanwu, Michael U},
  journal={BMC Infectious Diseases},
  volume={21},
  number={1},
  pages={975},
  year={2021},
  doi =  {https://doi.org/10.1186/s12879-021-06701-8}
  publisher={Springer}
}

@article{assiri2024unveiling,
  title={Unveiling the clinical spectrum of post-COVID-19 conditions: assessment and recommended strategies},
  author={Assiri, Abdullah M and Alamaa, Tareef and Elenezi, Faisal and Alsagheir, Aeshah and Alzubaidi, Lamya and TIeyjeh, Imad and Alhomod, Abdulazia S and Gaffas, Eisha M and Amer, Samar A and ELENEZI, Faisal and others},
  journal={Cureus},
  volume={16},
  number={1},
  year={2024},
  doi = {{https://doi.org/10.7759/cureus.52827}
  publisher={Cureus}
}

@article{he2025coinfection,
  title={Coinfection of COVID-19 and malaria: clinical profiles, interactions, and strategies for effective control},
  author={He, Mu-Zi and Zhang, Hai-Ting and Yang, Yi and Fang, Yi and Zhang, Mao and Deng, Sheng-Qun and Sun, Xun},
  journal={Malaria Journal},
  volume={24},
  number={1},
  pages={99},
  year={2025},
  doi = {https://doi.org/10.1186/s12936-025-05315-8}
  publisher={Springer}
}

@article{segbefia2024cytokine,
  title={Cytokine gene polymorphisms implicated in the pathogenesis of Plasmodium falciparum infection outcome},
  author={Segbefia, Selorm Philip and Asandem, Diana Asema and Amoah, Linda Eva and Kusi, Kwadwo Asamoah},
  journal={Frontiers in immunology},
  volume={15},
  pages={1285411},
  year={2024},
  doi = {https://doi.org/10.3389/fimmu.2024.1285411}
  publisher={Frontiers Media SA}
}

@article{bertoletti2022sars,
  title={SARS-CoV-2-specific T cells in the changing landscape of the COVID-19 pandemic},
  author={Bertoletti, Antonio and Le Bert, Nina and Tan, Anthony T},
  journal={Immunity},
  volume={55},
  number={10},
  pages={1764--1778},
  year={2022},
  doi = {https://doi.org/10.1016/j.immuni.2022.08.008}
  publisher={Elsevier}
}

@article{soma2022pathophysiological,
  title={Pathophysiological changes in erythrocytes contributing to complications of inflammation and coagulation in COVID-19},
  author={Soma, Prashilla and Bester, Janette},
  journal={Frontiers in Physiology},
  volume={13},
  pages={899629},
  year={2022},
  doi = {https://doi.org/10.3389/fphys.2022.899629}
  publisher={Frontiers Media SA}
}

@article{sinha2025power,
  title={Power of Memory: A Natural Killer Cell Perspective},
  author={Sinha, Oishi and Abhipsha, SK and Sen Santara, Sumit},
  journal={Cells},
  volume={14},
  number={11},
  pages={846},
  year={2025},
  doi ={https://doi.org/10.3390/cells14110846}
  publisher={MDPI}
}

@article{iesa2020sars,
  title={SARS-CoV-2 and Plasmodium falciparum common immunodominant regions may explain low COVID-19 incidence in the malaria-endemic belt},
  author={Iesa, MAM and Osman, MEM and Hassan, MA and Dirar, AIA and Abuzeid, N and Mancuso, JJ and Pandey, R and Mohammed, AA and Borad, MJ and Babiker, HM and others},
  journal={New microbes and new infections},
  volume={38},
  pages={100817},
  year={2020},
  doi = {https://doi.org/10.1016/j.nmni.2020.100817}
  publisher={Elsevier}
}

@article{tchoumi2021malaria,
  title={Malaria and COVID-19 co-dynamics: A mathematical model and optimal control},
  author={Tchoumi, SY and Diagne, ML and Rwezaura, H and Tchuenche, JM},
  journal={Applied mathematical modelling},
  volume={99},
  pages={294--327},
  year={2021},
  doi = {https://doi.org/10.1016/j.apm.2021.06.016}
  publisher={Elsevier}
}

@article{abioye2023fractional,
  title={A fractional-order mathematical model for malaria and COVID-19 co-infection dynamics},
  author={Abioye, Adesoye Idowu and Peter, Olumuyiwa James and Ogunseye, Hammed Abiodun and Oguntolu, Festus Abiodun and Ayoola, Tawakalt Abosede and Oladapo, Asimiyu Olalekan},
  journal={Healthcare Analytics},
  volume={4},
  pages={100210},
  year={2023},
  doi = {https://doi.org/10.1016/j.health.2023.100210}
  publisher={Elsevier}
}

@article{shah2022optimum,
  title={Optimum controls to avert co-infection of Malaria-COVID-19},
  author={Shah, Nita H and Sheoran, Nisha and Jayswal, Ekta N},
  journal={Journal of Statistics and Management Systems},
  volume={25},
  number={8},
  pages={2043--2059},
  year={2022},
  doi = {https://doi.org/10.1080/09720510.2021.2020429}
  publisher={Taylor \& Francis}
}

@misc{owid-covid-data,
  author       = {{Our World in Data}},
  title        = {Coronavirus Pandemic (COVID-19)},
  year         = {2020},
  url          = {https://ourworldindata.org/coronavirus},
  note         = {Accessed: 20 February 2026}
}
