## Supplementary material for "A Deterministic–Stochastic Model for COVID-19 and Malaria Co-Infection with Malaria-Acquired Partial Immunity": Latex: plos_latex_template.pdf

### Title of submission to PLOS journals

Name1 Surname[1,2◑], Name2 Surname[2◑], Name3 Surname[2,3¤], Name4 Surname[2], Name5 Surname[2‡], Name6 Surname[2‡], Name7 Surname[1,2,3*], with the Lorem Ipsum Consortium[¶]

**1** Affiliation Dept/Program/Center, Institution Name, City, State, Country
**2** Affiliation Dept/Program/Center, Institution Name, City, State, Country
**3** Affiliation Dept/Program/Center, Institution Name, City, State, Country

◑These authors contributed equally to this work.
‡These authors also contributed equally to this work.
¤Current Address: Dept/Program/Center, Institution Name, City, State, Country
†Deceased
¶Membership list can be found in the Acknowledgments section.
*

#### Abstract

Lorem ipsum dolor sit amet, consectetur adipiscing elit. Curabitur eget porta erat. Morbi consectetur est vel gravida pretium. Suspendisse ut dui eu ante cursus gravida non sed sem. Nullam sapien tellus, commodo id velit id, eleifend volutpat quam. Phasellus mauris velit, dapibus finibus elementum vel, pulvinar non tellus. Nunc pellentesque pretium diam, quis maximus dolor faucibus id. Nunc convallis sodales ante, ut ullamcorper est egestas vitae. Nam sit amet enim ultrices, ultrices elit pulvinar, volutpat risus.

#### Author summary

Lorem ipsum dolor sit amet, consectetur adipiscing elit. Curabitur eget porta erat. Morbi consectetur est vel gravida pretium. Suspendisse ut dui eu ante cursus gravida non sed sem. Nullam sapien tellus, commodo id velit id, eleifend volutpat quam. Phasellus mauris velit, dapibus finibus elementum vel, pulvinar non tellus. Nunc pellentesque pretium diam, quis maximus dolor faucibus id. Nunc convallis sodales ante, ut ullamcorper est egestas vitae. Nam sit amet enim ultrices, ultrices elit pulvinar, volutpat risus.

#### Introduction

Lorem ipsum dolor sit [1] amet, consectetur adipiscing elit. Curabitur eget porta erat. Morbi consectetur est vel gravida pretium. Suspendisse ut dui eu ante cursus gravida non sed sem. Nullam Eq (1) sapien tellus, commodo id velit id, eleifend volutpat quam. Phasellus mauris velit, dapibus finibus elementum vel, pulvinar non tellus. Nunc pellentesque pretium diam, quis maximus dolor faucibus id. [2] Nunc convallis sodales ante, ut ullamcorper est egestas vitae [3]. Nam sit amet enim ultrices, ultrices elit pulvinar, volutpat risus [4].

$$\mathrm{P_Y} = \underbrace{H(Y_n) - H(Y_n|\mathbf{V}_n^Y)}_{S_Y} + \underbrace{H(Y_n|\mathbf{V}_n^Y) - H(Y_n|\mathbf{V}_n^{X,Y})}_{T_{X\rightarrow Y}}, \tag{1}$$

### Materials and methods

#### Etiam eget sapien nibh

Nulla mi mi, Fig 1 venenatis sed ipsum varius, volutpat euismod diam. Proin rutrum vel massa non gravida. Quisque tempor sem et dignissim rutrum. Lorem ipsum dolor sit amet, consectetur adipiscing elit. Morbi at justo vitae nulla elementum commodo eu id massa. In vitae diam ac augue semper tincidunt eu ut eros. Fusce fringilla erat porttitor lectus cursus, S1 Video vel sagittis arcu lobortis. Aliquam in enim semper, aliquam massa id, cursus neque. Praesent faucibus semper libero.

**Fig 1. Bold the figure title.** Figure caption text here, please use this space for the figure panel descriptions instead of using subfigure commands. A: Lorem ipsum dolor sit amet. B: Consectetur adipiscing elit.

### Results

Nulla mi mi, venenatis sed ipsum varius, Table 1 volutpat euismod diam. Proin rutrum vel massa non gravida. Quisque tempor sem et dignissim rutrum. Lorem ipsum dolor sit amet, consectetur adipiscing elit. Morbi at justo vitae nulla elementum commodo eu id massa. In vitae diam ac augue semper tincidunt eu ut eros. Fusce fringilla erat porttitor lectus cursus, vel sagittis arcu lobortis. Aliquam in enim semper, aliquam massa id, cursus neque. Praesent faucibus semper libero.

**Table 1. Table caption Nulla mi mi, venenatis sed ipsum varius, volutpat euismod diam.**

| **Heading1** | | | | **Heading2** | | | |
|---|---|---|---|---|---|---|---|
| $cell1row1$ | cell2 row 1 | cell3 row 1 | cell4 row 1 | cell5 row 1 | cell6 row 1 | cell7 row 1 | cell8 row 1 |
| $cell1row2$ | cell2 row 2 | cell3 row 2 | cell4 row 2 | cell5 row 2 | cell6 row 2 | cell7 row 2 | cell8 row 2 |
| $cell1row3$ | cell2 row 3 | cell3 row 3 | cell4 row 3 | cell5 row 3 | cell6 row 3 | cell7 row 3 | cell8 row 3 |

Table notes Phasellus venenatis, tortor nec vestibulum mattis, massa tortor interdum felis, nec pellentesque metus tortor nec nisl. Ut ornare mauris tellus, vel dapibus arcu suscipit sed.

#### LOREM and IPSUM nunc blandit a tortor

##### 3rd level heading

Maecenas convallis mauris sit amet sem ultrices gravida [5]. Etiam eget sapien nibh. Sed ac ipsum eget enim egestas ullamcorper nec euismod ligula. Curabitur fringilla pulvinar lectus consectetur pellentesque. Quisque augue sem, tincidunt sit amet feugiat eget, ullamcorper sed velit [6]. Sed non aliquet felis. Lorem ipsum dolor sit amet, consectetur adipiscing elit. Mauris commodo justo ac dui pretium imperdiet. Sed suscipit iaculis mi at feugiat.

1. react
2. diffuse free particles
3. increment time by dt and go to 1

##### Sed ac quam id nisi malesuada congue

Nulla mi mi, venenatis sed ipsum varius, volutpat euismod diam. Proin rutrum vel massa non gravida. Quisque tempor sem et dignissim rutrum. Lorem ipsum dolor sit amet, consectetur adipiscing elit [7]. Morbi at justo vitae nulla elementum commodo eu id massa [8]. In vitae diam ac augue semper tincidunt eu ut eros. Fusce fringilla erat porttitor lectus cursus, vel sagittis arcu lobortis [9]. Aliquam in enim semper, aliquam massa id, cursus neque. Praesent faucibus semper libero.

- First bulleted item.
- Second bulleted item.
- Third bulleted item.

#### Discussion

Nulla mi mi, venenatis sed ipsum varius, Table 1 volutpat euismod diam [10]. Proin rutrum vel massa non gravida. Quisque tempor sem et dignissim rutrum. Lorem ipsum dolor sit amet, consectetur adipiscing elit. Morbi at justo vitae nulla elementum commodo eu id massa [11]. In vitae diam ac augue semper tincidunt eu ut eros. Fusce fringilla erat porttitor lectus cursus, vel sagittis arcu lobortis. Aliquam in enim semper, aliquam massa id, cursus neque. Praesent faucibus semper libero [12].

#### Conclusion

$CO_2$ Maecenas convallis mauris sit amet sem ultrices gravida. Etiam eget sapien nibh. Sed ac ipsum eget enim egestas ullamcorper nec euismod ligula. Curabitur fringilla pulvinar lectus consectetur pellentesque. Quisque augue sem, tincidunt sit amet feugiat eget, ullamcorper sed velit [13].

Sed non aliquet felis. Lorem ipsum dolor sit amet, consectetur adipiscing elit. Mauris commodo justo ac dui pretium imperdiet. Sed suscipit iaculis mi at feugiat. Ut neque ipsum, luctus id lacus ut, laoreet scelerisque urna. Phasellus venenatis, tortor nec vestibulum mattis, massa tortor interdum felis, nec pellentesque metus tortor nec nisl. Ut ornare mauris tellus, vel dapibus arcu suscipit sed. Nam condimentum sem eget mollis euismod. Nullam dui urna, gravida venenatis dui et, tincidunt sodales ex. Nunc est dui, sodales sed mauris nec, auctor sagittis leo. Aliquam tincidunt, ex in facilisis elementum, libero lectus luctus est, non vulputate nisl augue at dolor. For more information, see S1 Appendix.

#### Supporting information

**S1 Fig. Bold the title sentence.** Add descriptive text after the title of the item (optional).

**S2 Fig. Lorem ipsum.** Maecenas convallis mauris sit amet sem ultrices gravida. Etiam eget sapien nibh. Sed ac ipsum eget enim egestas ullamcorper nec euismod ligula. Curabitur fringilla pulvinar lectus consectetur pellentesque.

**S1 File. Lorem ipsum.** Maecenas convallis mauris sit amet sem ultrices gravida. Etiam eget sapien nibh. Sed ac ipsum eget enim egestas ullamcorper nec euismod ligula. Curabitur fringilla pulvinar lectus consectetur pellentesque.

**S1 Video. Lorem ipsum.** Maecenas convallis mauris sit amet sem ultrices gravida. Etiam eget sapien nibh. Sed ac ipsum eget enim egestas ullamcorper nec euismod ligula. Curabitur fringilla pulvinar lectus consectetur pellentesque.

**S1 Appendix. Lorem ipsum.** Maecenas convallis mauris sit amet sem ultrices gravida. Etiam eget sapien nibh. Sed ac ipsum eget enim egestas ullamcorper nec euismod ligula. Curabitur fringilla pulvinar lectus consectetur pellentesque.

**S1 Table. Lorem ipsum.** Maecenas convallis mauris sit amet sem ultrices gravida. Etiam eget sapien nibh. Sed ac ipsum eget enim egestas ullamcorper nec euismod ligula. Curabitur fringilla pulvinar lectus consectetur pellentesque.

#### Acknowledgments

This section is intended only for general acknowledgements and thanks. Any information related to funding, data availability, author contributions, etc. should be entered directly into their dedicated fields in the PLOS Editorial Manager submission system, which will then be incorporated into the appropriate section in your article during the production process.
