## Supplementary material for "A Deterministic–Stochastic Model for COVID-19 and Malaria Co-Infection with Malaria-Acquired Partial Immunity": Latex: README

This is Elsevier's new document class for typeset journal articles,
elsarticle.cls.  It is now accepted for submitted articles, both in
Elsevier's electronic submission system and elsewhere.

Elsevier's previous document class for typeset articles, elsart.cls,
is now over 10 years old. It has been replaced with this newly written
document class elsarticle.cls, which has been developed for Elsevier
by the leading TeX developer STM Document Engineering Pvt Ltd.

elsarticle.cls is based upon the standard LaTeX document class
article.cls. It uses natbib.sty for bibliographical references.

Bugs and problems with elsarticle.cls may be reported to the
developers of the class via.

The file manifest.txt provides a list of the files in the elsarticle
bundle.  The following are the main files available:
- elsarticle.dtx, the dtx file
- elsdoc.pdf, the user documentation
- elsarticle-template-num.tex, template file for numerical citations
- elsarticle-template-harv.tex, template file for name-year citations
- elsarticle-template-num-names.tex, template file for numerical 
  citations + new natbib option.  Eg. Jones et al. [21]
- elsarticle-num.bst, bibliographic style for numerical references
- elsarticle-harv.bst, bibliographic style for name-year references
- elsarticle-num-names.bst, bibliographic style for numerical 
  referencces + new natbib option for citations.

To extract elsarticle.cls from *.dtx: latex elsarticle.ins

The documentation file is elsdoc.tex in the contrib directory.  To
compile it:
1. pdflatex elsdoc
2. pdflatex elsdoc
3. pdflatex elsdoc

or

use the makefile. Use the target `all' (eg: make all).

The above procedure will create a print version, namely elsdoc.pdf.

Copyright 2007-2020, Elsevier. Bugs, feature requests, suggestions and
comments may be mailed to. elsarticle.cls,
related documentation and supporting packages are released under the
LATEX Project Public Licence, either version 1.3 or any later
version. This work has the LPPL maintenance status
'author-maintained'.
