## Supplementary figures and images for "A Deterministic–Stochastic Model for COVID-19 and Malaria Co-Infection with Malaria-Acquired Partial Immunity"

### AH6.png

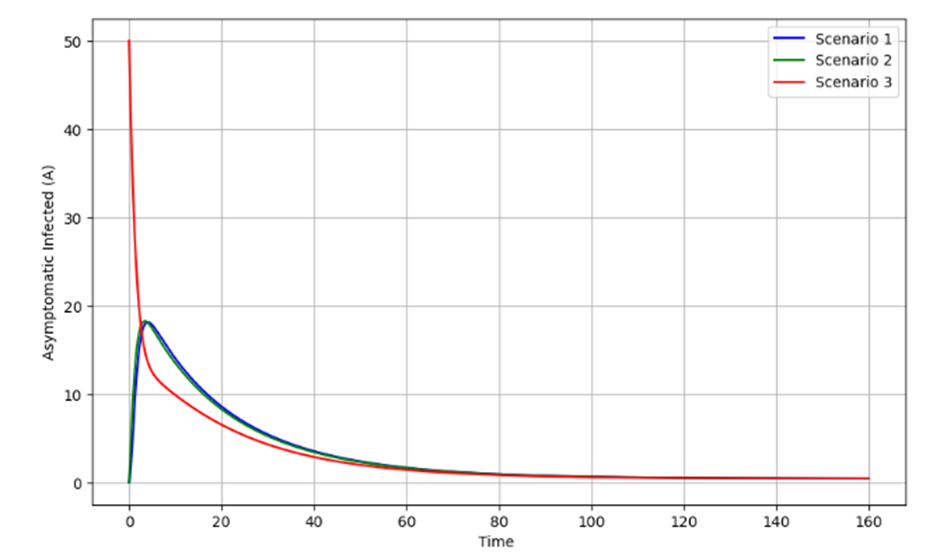

### COVID_MALARIA-4_page-0001.jpg

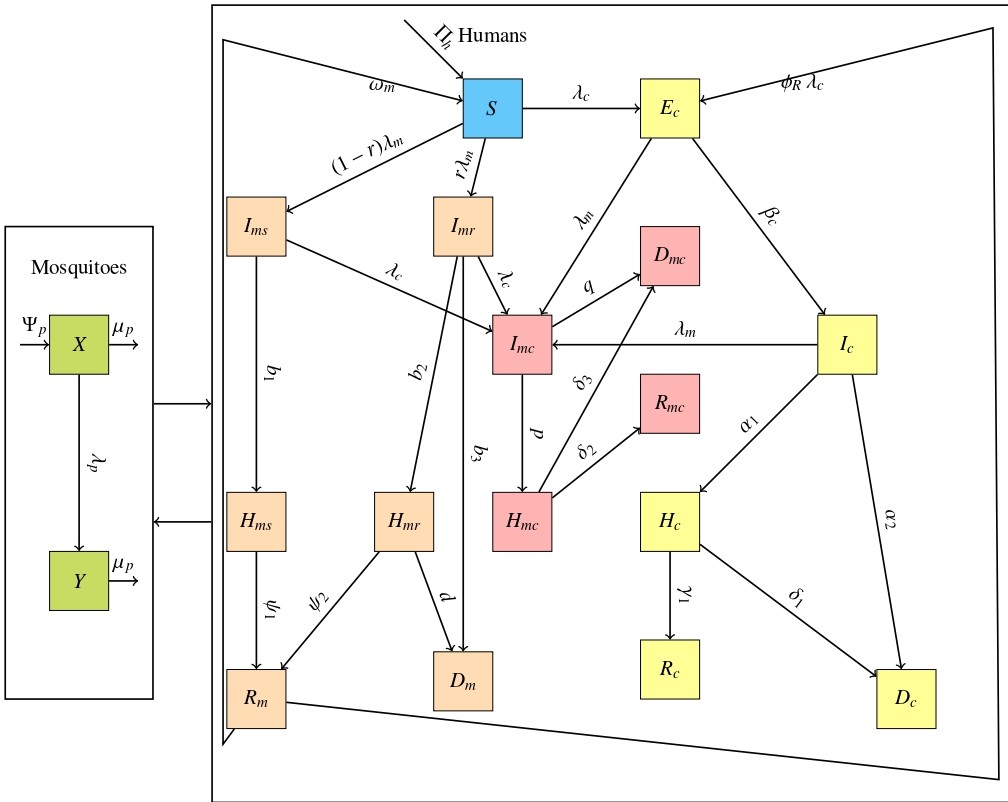

### covid_malaria_transparent_full.png

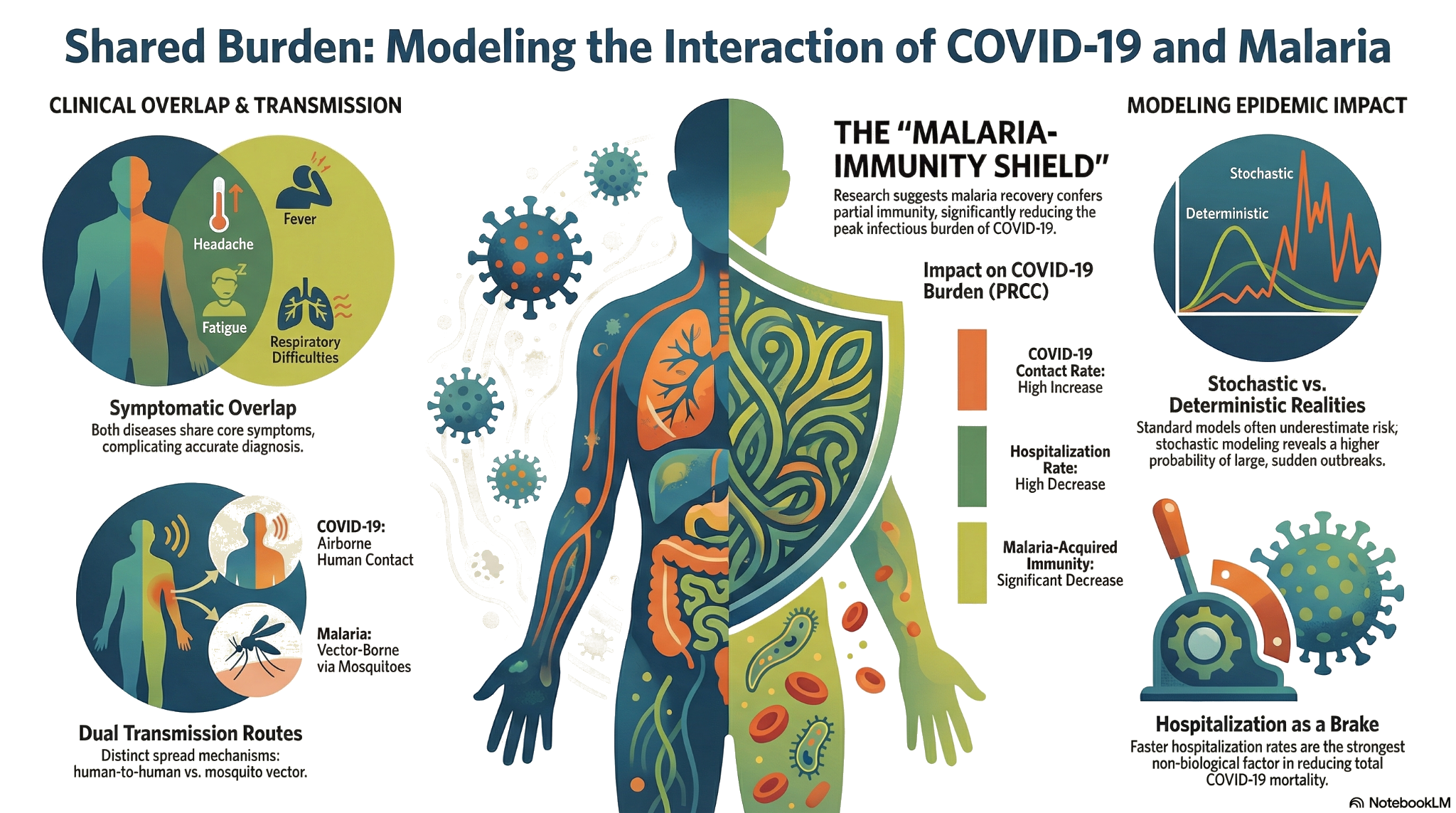

### imag2.png

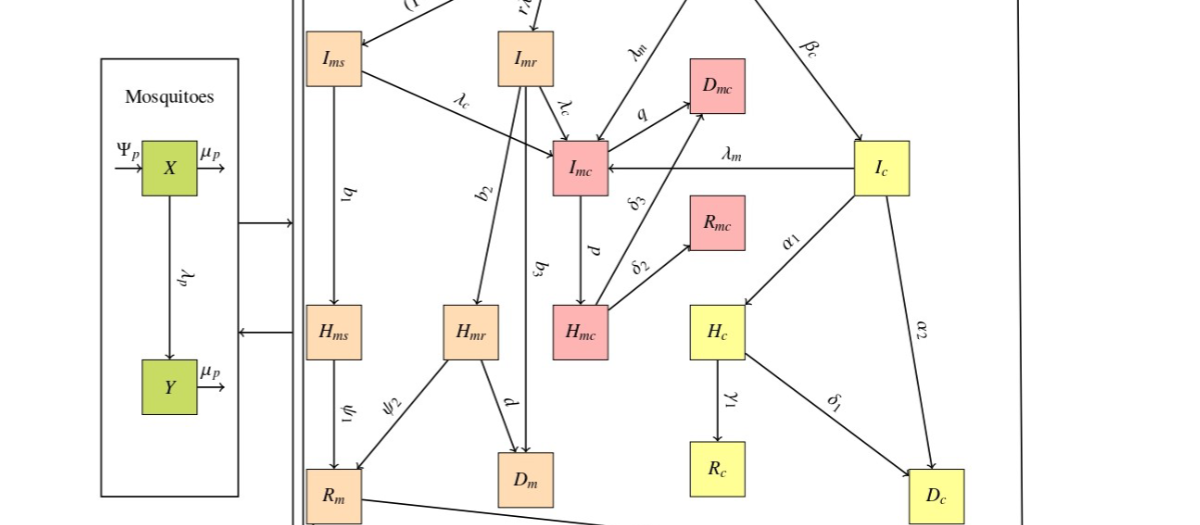

### image.png

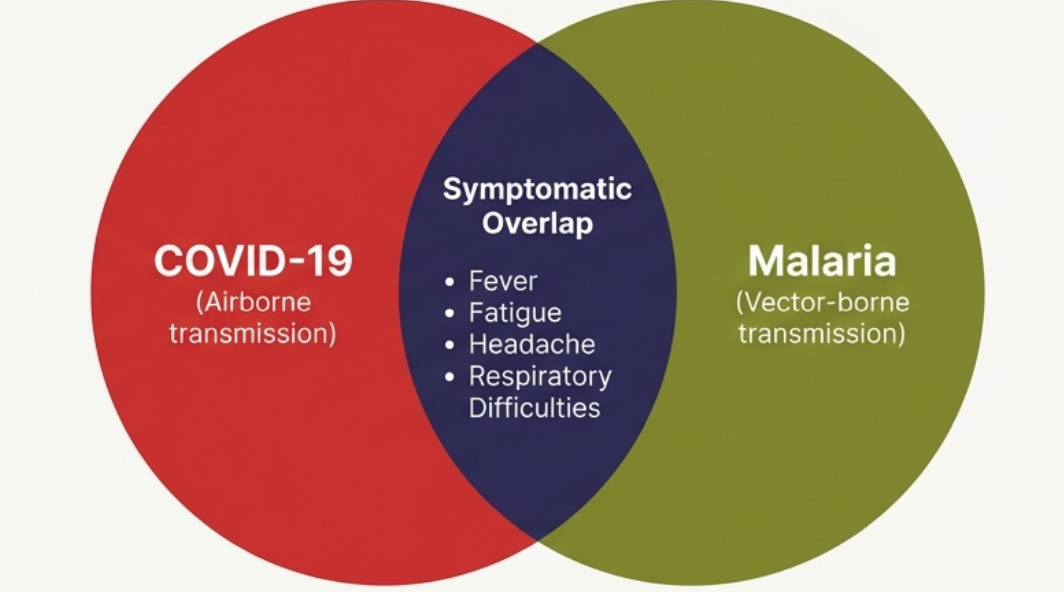
